# The Post-Septic Peripheral Myeloid Compartment Reveals Unexpected Diversity in Myeloid-Derived Suppressor Cells

**DOI:** 10.1101/2024.01.05.24300902

**Authors:** Evan L. Barrios, John Leary, Dijoia B. Darden, Jaimar C. Rincon, Micah Willis, Valerie E. Polcz, Gwendolyn S. Gillies, Jennifer A. Munley, Marvin L. Dirain, Ricardo Ungaro, Dina C. Nacionales, Marie-Pierre L. Gauthier, Shawn D. Larson, Laurence Morel, Tyler J. Loftus, Alicia M. Mohr, Robert Maile, Michael P. Kladde, Clayton E. Mathews, Maigan A. Brusko, Todd M. Brusko, Lyle L. Moldawer, Rhonda Bacher, Philip A. Efron

**Affiliations:** Sepsis and Critical Illness Research Center, Department of Surgery, University of Florida College of Medicine; Gainesville, Florida, USA; Department of Biostatistics, University of Florida College of Medicine and Public Health and Health Sciences, Gainesville, Florida, USA; Department of Biochemistry and Molecular Biology, University of Florida College of Medicine, Gainesville, Florida, USA; Department of Microbiology and Immunology, University of Texas San Antonio School of Medicine; San Antonio, Texas, USA; Department of Pathology, Immunology and Laboratory Medicine, University of Florida College of Medicine; Gainesville, Florida, USA

**Keywords:** myeloid-derived suppressor cells, sepsis, transcriptomics, single-cell RNA sequencing, chronic critical illness

## Abstract

Sepsis engenders distinct host immunologic changes that include the expansion of myeloid-derived suppressor cells (MDSCs). These cells play a physiologic role in tempering acute inflammatory responses but can persist in patients who develop chronic critical illness. The origins and lineage of these MDSC subpopulations were previously assumed to be discrete and unidirectional; however, these cells exhibit a dynamic phenotype with considerable plasticity. Using Cellular Indexing of Transcriptomes and Epitopes by Sequencing followed by transcriptomic analysis, we identify a unique lineage and differentiation pathway for MDSCs after sepsis and describe a novel MDSC subpopulation. Additionally, we report that the heterogeneous response of the myeloid compartment of blood to sepsis is dependent on clinical outcome.

## 1 Introduction

Sepsis is defined as life-threatening organ dysfunction caused by a dysregulated host response to infection (1), with survivors experiencing either a rapid recovery or chronic critical illness (CCI) (2). The emergency myelopoietic response to sepsis is characterized by hematopoietic stem and progenitor cell (HSPC) expansion and preferential differentiation along myeloid pathways (3-7). We and others have previously demonstrated that sepsis induces an expansion of circulating myeloid-derived suppressor cells (MDSCs), and that both an increase in and persistence of specific MDSC subpopulations are seen in sepsis patients with poor clinical outcomes (4, 8, 9).

Three MDSC subpopulations are typically described based on cell surface expression, mechanisms of immune suppression, and inflammatory profiles: granulocytic (PMN-), monocytic (M-), and early (E-) MDSCs (10, 11). Although these MDSCs differ phenotypically, they are all capable of suppressing T-lymphocyte proliferation (4, 12). As research into the myeloid compartment during inflammation expands, the complexity of intermediate cell types is just beginning to be understood. Indeed, Hegde et al. recently concluded that suppressive myeloid cell types, including MDSCs, *“are highly heterogeneous and context dependent”* (13). The authors presented an emergent view of MDSCs that emphasizes heterogeneity and plasticity in contrast to the classical view of MDSCs as the midpoint in a differentiation pathway that results in terminally-differentiated monocytes and granulocytes (13).

Single-cell RNA-sequencing (scRNA-seq) details the transcriptomes of complex and heterogeneous cell mixtures. An extension of this technique, Cellular Indexing of Transcriptomes and Epitopes by Sequencing (CITE-seq), simultaneously profiles cell surface proteins for each cell. We initially performed CITE-seq in order to identify MDSC subpopulations based on cell surface makers/cell phenotypes, as our previous results (14) indicated that MDSCs from septic patients may not express some of the classic genes found in MDSCs from cancer patients, making them difficult to identify. We compared the transcriptomes of myeloid cells from healthy subjects, acutely septic patients, and patients with good and poor clinical outcomes at later time points after sepsis. We found that MDSC subpopulations evolve over time and that outcome-dependent MDSC subpopulations exist. Specifically, we identified a novel hybrid (H)-MDSC phenotype unique to some sepsis survivors with poor clinical outcomes as well as acutely septic patients that progressed to CCI. Additionally, our findings suggest that the proliferation and cytokine production of lymphocytes, when co-cultured with MDSCs, vary at different time points after sepsis. Importantly, MDSCs do not express key genes seen in cancer whose downstream products suppress T-cell responsiveness. Overall, our results demonstrate a critical need for disease- or “context-” specific understanding of MDSCs when considering host-specific immune dysfunction and potential therapies.

## 2 Materials and Methods

### 2.1 Study Design

Our study design was previously reported by Darden, et al (14). To summarize, this prospective, observational cohort study was registered with *clinicaltrials.gov* (NCT02276417) and conducted at a tertiary care, academic research hospital. The objective of the study was to better understand the myeloid response (specifically blood MDSCs) to acute sepsis, and to identify transcriptomic differences in sepsis patients who rapidly recover versus those who develop CCI. Our hypothesis was that differences in the myeloid transcriptomic landscape could explain why some sepsis patients rapidly recover while others develop CCI.

Sepsis designation occurred through an electronic medical record-based Modified Early Warning Signs-Sepsis Recognition System (MEWS-SRS), which uses white blood cell count, heart rate, respiration rate, blood pressure, and mental status to identify patients at-risk for sepsis. All patients with sepsis were treated with early goal-directed fluid administration, initiation of broad-spectrum antibiotics, and vasopressor administration if appropriate. Antibiotic treatment was tailored to culture results and antibiotic resistance information.

Inclusion criteria:

Admission to the intensive care unit (ICU)
Age >17 years
Diagnosis of sepsis or septic shock according to the 2016 SCM/ESICM
International Sepsis Definitions Conference (1)
Initial septic episode while hospitalized
Management of patient via the sepsis clinical management protocol (15)

Exclusion criteria:

Refractory shock
Inability to achieve source control
Pre-sepsis expected lifespan <3 months
Expected withdrawal of care
Severe congestive heart failure (NYHA Class IV)
Child-Pugh Class C liver disease or undergoing evaluation for liver transplant
HIV infection with CD4^+^ count <200 cells/mm^3^
Prior organ transplant, use of chronic steroids, or immunosuppressive agents
Pregnancy
Institutionalized or other vulnerable patients
Chemotherapy or radiotherapy treatment within 30 days of sepsis onset
Severe traumatic brain injury (defined by radiologic evidence and GCS <8)
Spinal cord injury with permanent deficits
Unable to obtain informed consent

CCI was defined as ICU length of stay >13 days with persistent organ dysfunction as measured by the Sequential Organ Failure Assessment (SOFA) Score. Patients were also designated CCI with <14 days ICU length of stay if they were transferred to another hospital, or discharged to a long-term acute care facility or hospice with evidence of persistent organ dysfunction (4, 16). Patients were excluded from analysis if they died within 14 days of onset of sepsis (4, 16).

### 2.2 Human blood collection and sample preparation

Whole blood samples were collected at the following time points: 4 ± 1 days and 14-21 days after sepsis (16). For the former, we enrolled four patients diagnosed with septic shock (1) in order to guarantee a strong host response and transcriptomic alterations in circulating leukocytes. Interestingly, only half of these septic shock patients went on to develop CCI; the mortality for this cohort was 50% (17, 18). Samples from five patients who developed CCI and four patients who rapidly recovered after sepsis were obtained from additional patients between days 14-21 after sepsis diagnosis. We previously determined this time point to be key to MDSC differentiation as well as distinguishing CCI from rapid recovery (4, 16). In addition, whole blood was collected from 12 healthy subjects (1, 2). The proportion of men and women did not differ between sepsis patients and healthy subjects. Healthy subjects trended towards being younger than sepsis patients, although they still met the criteria of middle age (>45 years), encompassing patients who have poor outcomes after sepsis compared to younger cohorts (17, 18). Healthy subjects and septic patients had similar comorbidity scores, and similar underlying comorbidities (most commonly hypertension, chronic obstructive pulmonary disease, and diabetes mellitus).

Each blood collection underwent two enrichment procedures. PBMCs were collected from half of each sample using Ficoll-Paque^TM^ PLUS (Millipore Sigma, St. Louis, MO) and density gradient centrifugation. Myeloid cells were collected using RosetteSep^TM^ HLA Myeloid Cell Enrichment Kit (STEMCELL Technologies, Vancouver). A 1:3 mixture of enriched PBMCs: myeloid cells was created in order to adequately analyze the small target population (MDSCs, especially in control subjects) while also allowing for characterization of other important immune cell populations using CITE-seq.

#### 2.2.1 Human T-cell isolation and proliferation assay

Total T cells in the PBMC suspension were captured by immunomagnetic negative selection using EasySep^TM^ Human T Cell Isolation Kit (STEMCELL Technologies, Vancouver) according to the manufacturer’s instructions. Isolated CD3^+^ lymphocytes were labeled with cell trace violet (Thermo Fisher, Waltham, MA) to assess T-cell proliferation. T lymphocytes (1 x 10^5^ CD3^+^) were seeded into a 96-well plate and stimulated with soluble anti-CD3/CD28 antibodies (STEMCELL Technologies, Vancouver) or without, which served as the control. CD66b^+^ cells were also isolated from the PBMC fractions using EasySep^TM^ positive selection kit (STEMCELL Technologies, Vancouver) and were co-cultured with stimulated T cells in a 1:1 ratio at 37°C and 5% CO_2_. After 4 days, cells were harvested and supernatants were obtained for cytokine analysis. Cells were stained with anti-CD8 FITC and anti-CD4 PE and analyzed via flow cytometry (ZE5 Cell Analyzer, Bio-Rad Laboratories, CA). Proliferation indices were calculated as the total number of cell divisions divided by the number of cells that went into division (considering cells that underwent at least one division).

#### 2.2.2 Cytokine analysis

Human high sensitivity T cell magnetic bead 6-plex panels (IFN-γ, IL-10, IL-12 (p70), IL-17α, IL-2, IL-23) were purchased from EMD Millipore (Billerica, MA). Supernatants after cell culture were used for T cell-associated cytokines. The xPONENT software (EMD Millipore, Billerica, MA) was used for cytokine analysis.

#### 2.2.3 Flow cytometry

PBMC samples were analyzed fresh (not frozen and rethawed) due to differential viability of cell populations, particularly granulocytes.(19, 20) Although the PBMC fraction excludes mature granulocytes, it does contain low-density granulocytes that are presumed to include PMN-MDSCs (21). Classically, human blood MDSCs are defined from PBMCs as: M-MDSCs (CD11b^+^CD14^+^CD33^+^HLA^-^DR^low/-^) and PMN-MDSCs (CD11b^+^CD15^+^HLA^-^ DR^low^CD66b^+^) (22). Our preliminary flow cytometric analysis revealed that CD15 was not a good cell surface marker to isolate CD14^-^ cells from PBMCs (**Fig. S1C**). In fact, the analysis of CD33^+^CD11b^+^HLA^-^DR^low/-^ cells revealed heterogeneity in CD14 and CD15 cell surface expression. Thus, we chose to isolate CD66b^+^ cells from PBMCs to obtain PMN-MDSCs (**Fig. S1C**).

### 2.3 Statistics

#### 2.3.1 scRNA-seq read preprocessing

Gene expression and feature-barcoding data were generated using 10x Genomics v1.1 5’ chemistry and were sequenced on an Illumina HiSeq® with a target of 10,000 cells per sample (23). The **Cell Ranger** (10X Genomics) software suite was used to process base calls into FASTQ files, which were checked for quality control aberrations using **FastQC** v0.11.7 (24). A spliced + intronic, or *6plice*, reference transcriptome was generated from the hg38 reference genome (25). Reads were pseudoaligned to the reference transcriptome with **alevin-fry** v 0.8.1; USA mode was used for gene expression reads in order to provide separate quantifications of spliced, unspliced, and ambiguous mRNA abundance (26-28). The counts of 11 cell surface proteins of interest were also quantified using **alevin-fry**. Splicing-aware gene expression quantification was performed using Ensembl transcript IDs, with final counts matrices aggregated using Ensembl gene IDs.

#### 2.3.2 scRNA-seq data processing

Downstream data processing and analysis were performed primarily in R v4.2.3, with some additional processes written in Python v3.8 as required (29, 30). After loading the unfiltered spliced, unspliced, and ambiguous mRNA counts into R using **fishpond** package v2.4.1, we defined total mRNA counts as the elementwise sum of all three counts matrices and added the ambiguous counts to the spliced counts matrix (31). Unless otherwise specified, total mRNA counts were used as input throughout the analysis. Empty droplets and ambient mRNA were then identified and filtered out using the **DropletUtils** package v1.18.1 (32, 33). Cells with an estimated false discovery rate of <0.01 were kept for each sample. Next, the percentage of spliced reads coming from mitochondrial genes was computed for each cell, and cells with less than 5% mitochondrial DNA were kept (no significant difference between healthy and septic samples). Cell surface protein counts were imported as well, and cells that had valid gene expression barcodes but not protein barcodes were assigned a value of “0” for each protein. The raw counts matrices were then formatted and merged using the **Seurat** package v4.3.0, providing a single object with total, unspliced, and spliced mRNA as well as cell surface protein assays (34). Cells with less than three spliced and unspliced transcripts were removed by filtering; thus, the final merged dataset comprised 28,952 genes and 119,062 cells.

The total mRNA counts were scaled by library size factors and log1p-normalized, while protein counts were normalized via a centered log ratio transformation across each gene. Four thousand highly variable genes (HVGs) were identified using a local polynomial regression between the log of expression variance and the log of mean expression as implemented in the **FindVariableFeatures** function. After scaling the normalized counts, 100 principal components were computed using the set of HVGs as input, and each cell was scored and assigned a cell cycle phase as described previously (35). Next, the 25 different samples were integrated by the **Harmony** package v0.1.1, which corrects the existing Principal Component Analysis (PCA) embedding (36). The first 50 principle components were used as input, and a two-dimensional UMAP embedding was computed using the cosine distance on the resulting 50 Harmony components (37). Lastly, an approximate shared nearest neighbors graph was computed on the first 50 Harmony components using the cosine distance with the number of nearest neighbors set to 100, and the resulting graph was partitioned into clusters via Louvain modularity optimization using a resolution of 0.1 (38).

#### 2.3.3 scRNA-seq annotation

After clustering, the **SingleR** package v2.0.0 was used with several different immune reference datasets with known labels to assign a most-likely broad cell type to each cluster (39-44). In addition, the **Azimuth** package was used to map reference labels from an annotated dataset of healthy human PBMCs to each cell at multiple levels of granularity (34). Lastly, between-cluster differential expression testing was performed using the Wilcoxon rank-sum test with *p*-values adjusted via the Bonferroni correction. Genes were considered for testing if they were expressed by at least 25% of the cells in the cluster being tested, and results were retained if the mean log2 fold-change was greater than 0.25 and the adjusted *p*-value was less than 0.05 (45, 46). After a comparison of the resulting differentially expressed gene sets (DEGs) with canonical marker genes from the literature and an investigation of the unsupervised annotations, a broad cell type identity was manually assigned to each cluster.

After subsetting the initial dataset to just the cluster labeled as monocytes, cells with confident T-cell labels from Azimuth were filtered out and the data were split into two groups based on whether the cells came from healthy subjects or septic patients. Subcluster analysis was performed on the monocytes from the healthy subjects and septic patients. Briefly, the data were reprocessed and reintegrated as described before, though the number of HVGs was lowered to 3,000 and only 30 principal components were used as input to the integration, nonlinear dimension reduction, and clustering routines. In addition, the number of estimated nearest neighbors was reduced based on the smaller sizes of the subsets. Any further subclustering of heterogeneous cell types was performed using the same methods. Differential expression testing was again used to identify potential marker gene sets, and a fine cell type label was manually assigned to each cluster. Lastly, the cell type labels were subjected to confirmatory analysis using the available cell surface protein data as needed.

#### 2.3.4 scRNA-seq differential expression

Differential expression testing between for each time point versus healthy subjects in the “*classically*” annotated MDSCs was performed using a pseudobulk approach. Counts across all cells for each patient were aggregated and summed, then the DESeq2 method was applied for differential testing using the muscat R package v1.14.0 (47). The cell-type specific marker gene expression testing on the MDSCs annotated using the “*emergent*” view was performed using the **FindAllMarkers** function in Seurat using the wilcox method.

Differentially expressed testing between sepsis groups was performed using linear mixed models. Each gene was tested between comparison groups for M-MDSCs, as they were the only population of MDSCs with a sufficient number of cells (n > 50) per group. Normalized expression was used as the response, with a binary indicator for sepsis group as the sole fixed effect. A random intercept was included for each sample, and models were fit via the maximum likelihood estimation using the **MixedModels.jl** Julia package (48, 49). After recording expression statistics such as mean expression per group, raw fold change, and log2 fold change, the *p*-value of the group difference fixed effect from the linear mixed model was used to determine the significance of differential expression after adjustment using the Holm correction (38, 50).

#### 2.3.5 Enrichment of genes with high transcriptional activity in MDSCs

The unspliced ratio per gene per cell was calculated as (unspliced counts + 1) divided by the (spliced counts + 1), then the mean for each gene was calculated separately for the MDSC subpopulations. Genes having a mean ratio greater than 1.1 were considered as having a high degree of active transcription. The gprofiler2 R package v0.2.1 (51) was used to identify significantly enriched biological processes for each set of genes, then a network-based approach was performed to better understand the biological functions using the **vissE** R package v1.8.0 (52). Similarities among the enriched processes were computed using the Jaccard index and then used to build an overlap network. Clusters of enriched gene-sets were identified by graph clustering; for each cluster a frequency analysis of words in the gene-set names indicates the most relevant biological functions.

#### 2.3.6 scRNA-seq trajectory inference and RNA velocity

After annotating the septic monocytic cells, the data were further subject to only include the cell types thought to be relevant to MDSC development and differentiation: classical and non-classical monocytes, cDCs, and MDSCs. This subset was re-embedded using UMAP, and the cells were reclustered using the re-computed simulated neural network graph as input to the Louvain algorithm (37). After extracting the UMAP parameters from the output of the **RunUMAP** function, we used the **uwot** R package to regenerate the fitted UMAP model and nearest neighbor data that were generated internally (53). From this output we extracted the UMAP connectivity graph, which is a sparse representation of the fuzzy simplicial data set that can be loosely interpreted as a metric of how likely connections are between cells (37). The raw counts matrices, metadata for cells and genes, nearest neighbor graphs, PCA, Harmony, and UMAP embeddings, and the UMAP connectivity graph were used to generate an **AnnData** object in Python that exactly matched the preprocessing used when annotating the cells in R (54).

The preprocessed data were used as input to an RNA velocity estimation workflow built around the **scVelo** package v0.2.5. After computing first-order moments of the spliced and unspliced counts, the dynamical velocity model was used to estimate per-gene velocities and a cell-level velocity graph, after which the velocities were projected onto the existing UMAP embedding (55, 56). Next, transition probability matrices, absorption probabilities, and initial and terminal cell state likelihoods were estimated based on a weighted kernel of the velocity estimates and UMAP connectivities using the **CellRank** package v1.5.1 (57). The resulting cell fate probabilities then served as a prior for the estimation of a gene-shared latent time for each cell. Lineage driving genes were identified by estimating the Spearman correlation of each gene’s expression with absorption probabilities for each identified cell fate. Finally, a directed partitioned graph abstraction was estimated and projected into the existing UMAP embedding using the state probability and latent time estimates as priors; these computations were performed using the partition-based graph abstraction (PAGA) algorithm as implemented in v1.9.3 of the **Scanpy** package (58, 59). In addition, an undirected graph abstraction was used as the initialization for a force-directed graph embedding of the cells, after which the graph abstraction was recomputed on the resulting embedding. This layout of the cells was used to display inter-cell type connectivities, which were estimated as described previously using UMAP (60, 61).

Differences in the dynamical model parameters (state probabilities, velocity length and pseudotime, cell stability index, and lineage priming) were tested between septic groups within MDSC subpopulations using a linear mixed model. Specifically, the **nlme** R package v3.1-162 (62) was used to fit a model with fixed effects of cell type, group, and their interaction, and a random intercept for subject. Pairwise testing was then obtained using contrasts of interest (across groups within cell type) with the **emmeans** R package v1.8.7 (63).

## 3. Results

### 3.1 MDSC subpopulations initially defined by classical cell surface markers

Here we have used CITE-seq to analyze single-cell transcriptomic profiles of MDSCs in blood from healthy subjects (n=12) and surgical sepsis patients at 4 ± 1 (n=4) and 14-21 (n=9) days after sepsis onset (64). Septic patients at 14-21 days were further divided based on their clinical outcomes at time of sampling, defined as either ‘rapid recovery’ (n=4) or development of CCI (n=5). CCI was defined as sepsis survivors requiring 14 or more days of ICU care with persistent organ injury. Sex, age, BMI, and comorbidity profiles were similar between cohorts (**Table 1**).

**Table 1.** Patient characteristics between cohorts. Cohorts are healthy control patients, acutely septic patients, and late sepsis patients who experienced rapid recovery (RAP) and chronic critical illness (CCI). BMI: body mass index, CCI: Charlson comorbidity index, COPD: chronic obstructive pulmonary disease, DM: diabetes mellitus, HTN: hypertension, NSTI: necrotizing soft tissue infection, SBO: small bowel obstruction, MCC: Motorcycle crash.

|  | Healthy Subjects<br>(n=12) | Sepsis Day 4 ± 1<br>(n=4) | RAP Days 14-21<br>(n=4) | CCI Days 14-21<br>(n=5) | p-value |
| --- | --- | --- | --- | --- | --- |
| Male, # (%) | 7 (58) | 1 (25) | 1 (25) | 3 (60) | 0.48 |
| Age in years, (μ ± SD) | 46 ± 10 | 67 ± 22 | 61 ± 16 | 58 ± 18 | 0.08 |
| BMI (μ ± SD) |  | 39 ± 19 | 37 ± 20 | 21 ± 3 | 0.19 |
| Septic shock, # (%) |  | 4 (100) | 1 (25) | 4 (80) |  |
| CCI (median) |  | 5.5 | 2 | 2 |  |
| Comorbidities (#) | Cancer (1), COPD (1), DM (1), HTN (3) | COPD (1), DM (2), HTN (4) | COPD (1), DM (1), HTN (4) | DM (1), HTN (2) |  |
| Admission Diagnosis (#) |  | NSTI (1), Choledocholithiasis (1), SBO (1), Planned operation (1) | NSTI (2), SBO (2) | Planned operation (1), Complication (1), Intra-abdominal abscess (1), Pancreatitis (1), MCC (1) |  |

Similar to flow cytometry phenotyping, CITE-seq employs conventional cell surface markers for myeloid cell subpopulations to define E-MDSCs (Lin^−^HLADR^low/−^ CD33^+^CD11b^+^CD14^−^CD15^−^CD66b^−^), PMN-MDSCs (Lin^−^CD33^+^CD11b^+^CD14^−^ and CD15^+^ or CD66b^+^), and M-MDSCs (Lin^−^HLADR^low/−^CD33^+^CD11b^+^CD14^+^CD15^−^CD66b^−^), as well as CD14^+^CD16^−^ (classical) and CD14^dim^CD16^+^ (non-classical) monocytes (while removing platelets, erythrocytes, HSPCs, γδ T cells, and innate lymphoid cells). This is consistent with the classical monolithic view of myeloid differentiation described by Hegde (**Fig. 1A**) (13). Historically, flow cytometry classification of MDSCs is performed directly on isolated PBMCs (4, 16), and our analysis revealed that PMN-MDSCs made up the majority of MDSCs in isolated PBMCs of representative septic patients, consistent with prior literature (**Table 2**) (4, 16).

**Figure 1.**
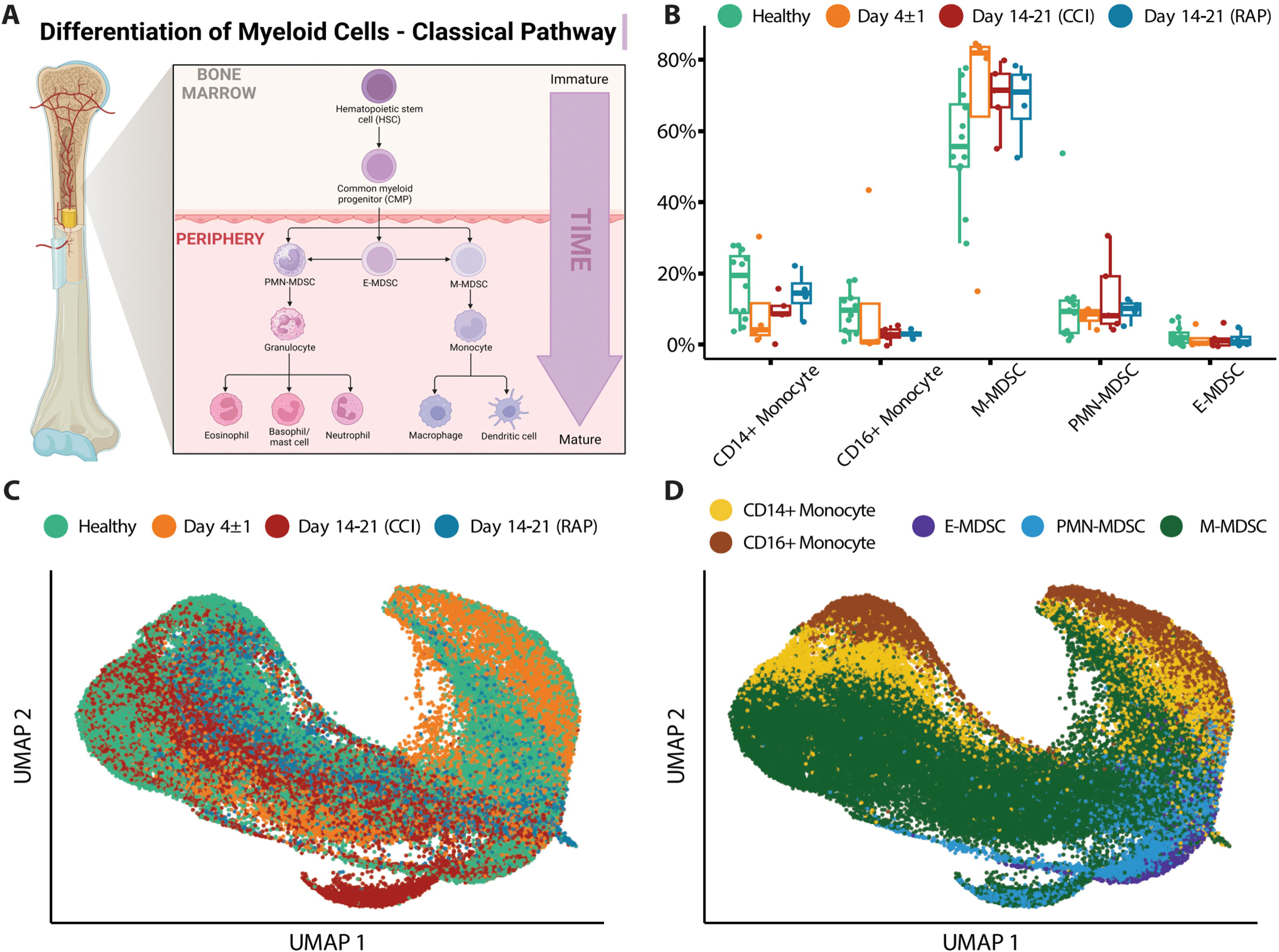
Single-cell analysis of myeloid cells using surface protein makers. **(A)** Illustration representing the historical/classic/monolithic definition of MDSCs. E-, PMN-, and M-MDSCs are the predominant subpopulations with distinct phenotypes and functions (modified from Hegde et al. *(13)*). **(B)** Cell proportions of monocyte subtypes and MDSCs relative to overall monocytic cells are shown for healthy subjects (“Healthy”) (n=12), septic patients 4 days following diagnosis (“Day 4 ± 1”) (n=4), and septic patients at days 14-21 (separated into those experiencing chronic critical illness (“CCI”) (n=5) or those who rapidly recovered (“RAP”) (n=4)). **(C)** UMAP embedding of single-cell transcriptomes of peripheral blood mononuclear cells (PBMCs). Cells are colored by the timepoint at which the samples were taken. Samples from day 4 and days 14-21 are from septic patients. **(D)** Similar to **(C)**, with cells colored by cell type. M: monocytic, PMN: granulocytic, E: early.

**Table 2.** Percentage of Total MDSCs and MDSC subpopulations from PBMCs via flow cytometry. Percentages of total MDSC population in representative septic patients. Blood was collected from healthy subjects (n=6), day 4 ± 1 septic patients (n=7), and late sepsis patients at days 14-21 (n=3)). PBMCs were isolated and prepared for flow cytometry. Viable cells determined followed by gating of CD11b^+^ and CD33^+^ cells. HLA-DR^low^ cells selected to capture Total MDSCs. (CD11b^+^ CD33^+^ HLA-DR^low^) Cells outside the gating of the three MDSC subpopulations are classified as “% Ungated.”

| MDSC Subpopulation | Healthy Subjects (n=6) | Sepsis Day 4 ± 1 (n=7) | Sepsis Days 14-21 (n=3) |
| --- | --- | --- | --- |
| % Total MDSCs | 15.1 (8.1, 16.6) | 39.2 (25.2, 55.6) | 44.8 (35.6, 56.3) |
| % E-MDSC | 68.1 (49.8, 78.7) | 1.4 (0.7, 5.9) | 2.3 (2.2, 9.0) |
| % PMN-MDSC | 14.6 (7.4, 36.9) | 79.5 (64.4, 89.0) | 80.7 (52.0, 85.1) |
| % M-MDSC | 9.2 (4.9, 12.7) | 9.1 (8.1, 12.7) | 11.5 (9.7, 35.9) |
| % Ungated | 2.0 (1.4, 3.2) | 0.7 (0.5, 1.3) | 0.7 (0.7, 3.1) |

We then evaluated cell proportions using transcriptomics with confirmation via flow cytometry. Myeloid cell enrichment was necessary in this step in order to detect MDSCs in healthy subject samples during CITE-seq as they are a relatively rare population (*see Methods Section entitled “Human Blood Collection and Sample Preparation”*). Both single-cell transcriptomics and flow cytometry revealed an overall increase in total MDSCs acutely after sepsis (**Fig. 1B and Table 3**). We plotted the cells via Uniform Manifold Approximation and Projection (UMAP) based on timepoint after sepsis (**Fig. 1C**) and myeloid cell subtype (**Fig. 1D**), which revealed heterogeneity of these cells when analyzing their single cell transcriptomes. The classification of MDSCs based on cell surface phenotypes is somewhat dependent on the method of analysis, and as the myeloid enrichment kit (STEMCELL) uses CD33 (and CD33 is expressed on all MDSCs), this was our first inclination that an alternative method of classifying cells by subpopulation would be necessary.

**Table 3.**
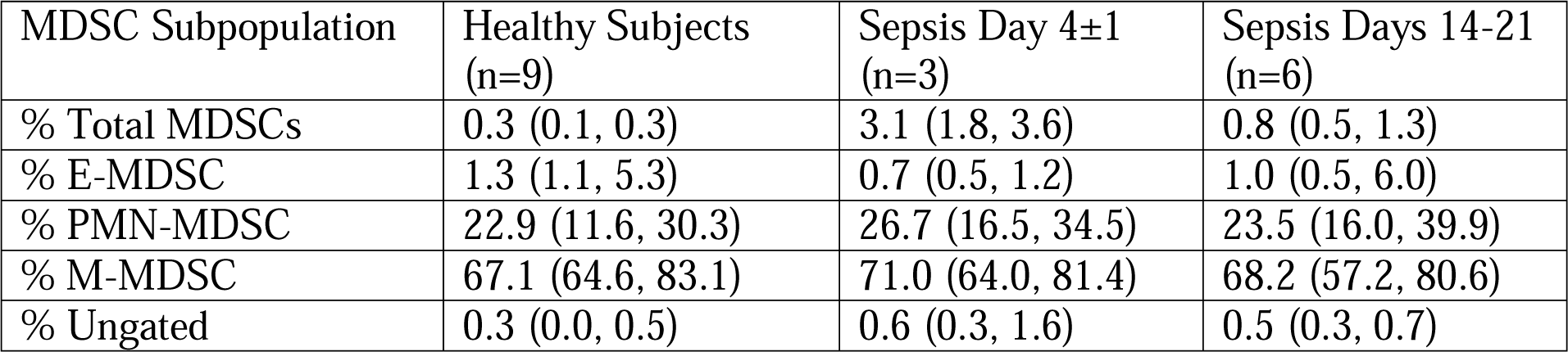
Percentage of Total MDSCs and MDSC subpopulations from PBMCs and enriched myeloid cells via flow cytometry. Percentages of total MDSC population in representative septic patients. Peripheral blood mononuclear cells and myeloid cells were collected from the same septic cohorts (n=3 for acute sepsis and n=6 for late sepsis patients) and healthy subjects (n=9). A 3:1 mixture of myeloid cells: enriched PBMCs were prepared for flow cytometry. Cells outside the gating of the three MDSC subpopulations are classified as “% Ungated.” Results reported as median (Q1, Q3). E: early, PMN: granulocytic, M: monocytic.

| MDSC Subpopulation | Healthy Subjects (n=9) | Sepsis Day 4±1 (n=3) | Sepsis Days 14-21 (n=6) |
| --- | --- | --- | --- |
| % Total MDSCs | 0.3 (0.1, 0.3) | 3.1 (1.8, 3.6) | 0.8 (0.5, 1.3) |
| % E-MDSC | 1.3 (1.1, 5.3) | 0.7 (0.5, 1.2) | 1.0 (0.5, 6.0) |
| % PMN-MDSC | 22.9 (11.6, 30.3) | 26.7 (16.5, 34.5) | 23.5 (16.0, 39.9) |
| % M-MDSC | 67.1 (64.6, 83.1) | 71.0 (64.0, 81.4) | 68.2 (57.2, 80.6) |
| % Ungated | 0.3 (0.0, 0.5) | 0.6 (0.3, 1.6) | 0.5 (0.3, 0.7) |

Next, we performed pseudobulk differential gene expression between septic patients at day 4 and days 14-21 post-sepsis diagnosis compared to healthy subjects to assess possible differences among septic groups. Dramatic differences in gene expression within MDSC subpopulations (specifically PMN- and M-MDSCs, which were most abundant) were observed that varied over time (**Fig. 2**). Considering PMN-MDSCs, by comparing each differential expression test performed against healthy subjects, we found more extreme fold-changes in late sepsis patients who developed CCI compared to acutely septic patients (**Fig. 2A, left panel**). Conversely, gene expression for late sepsis patients who rapidly recovered returned towards that seen in healthy subjects when compared to both acute sepsis and late sepsis with CCI. Gene expression for rapid recovery and CCI patients compared to healthy subjects also tended to diverge (**Fig. 2A, middle and right panels**). Overall, 52 genes were differentially expressed in PMN-MDSCs from acutely septic patients; however, only three of these genes were also significantly differentially expressed in both late sepsis patients who either rapidly recovered or developed CCI (**Fig. 2B; Supplemental File 1**). The ontology of transcriptional differences among septic patients at different time points also illustrated the heterogeneity of the PMN-MDSC response over time (**Fig. 2C**).

**Figure 2.**
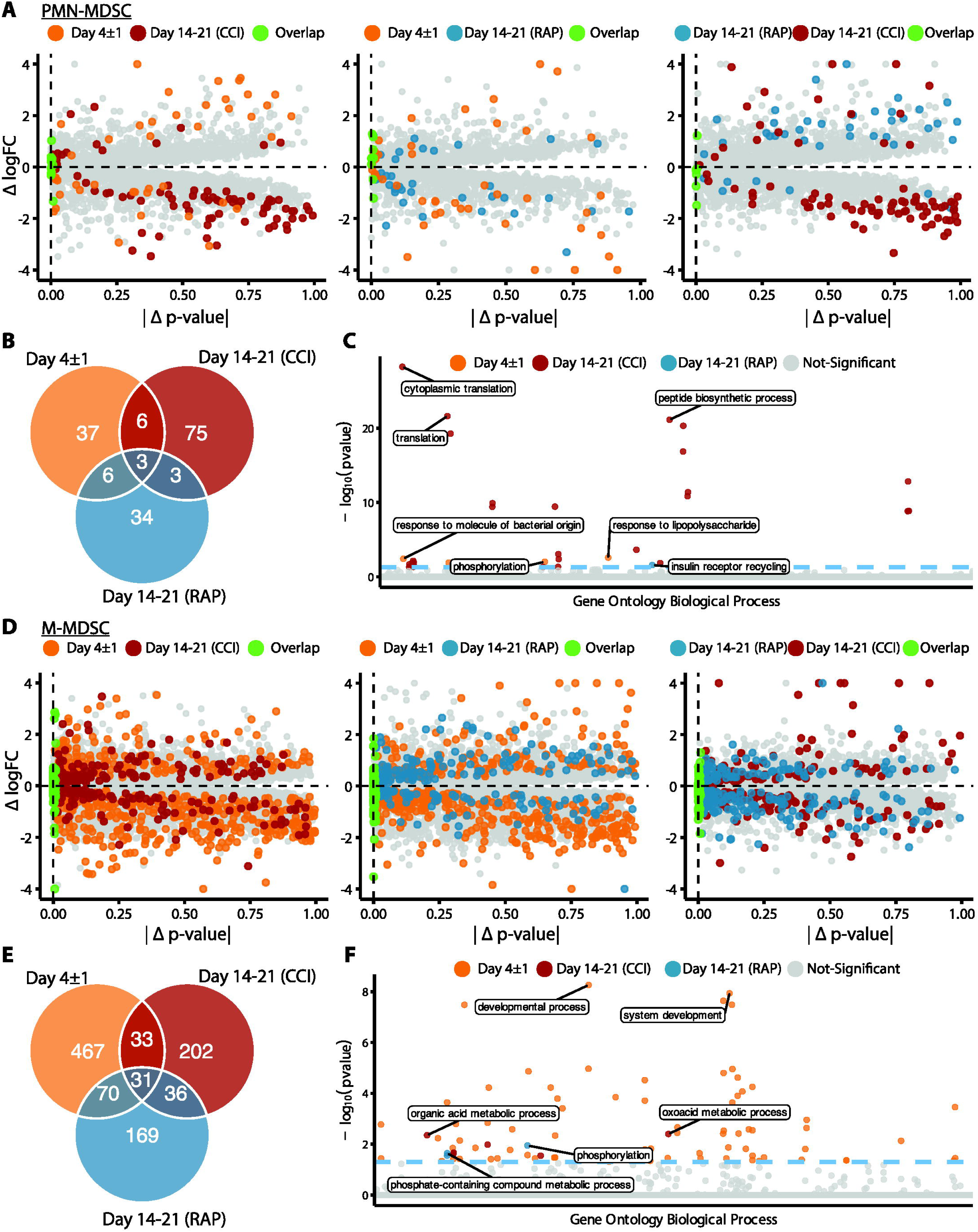
Analysis via CITE-seq of differential gene expression of PMN- and M-MDSC subpopulations at different time points relative to healthy subjects. **(A)** Within PMN-MDSCs, gene expression of twelve healthy subjects (baseline) was compared with septic patients at day 4 (“Day 4 ± 1”) (n=4) and septic patients at days 14-21 (subdivided into chronic critical illness (“CCI”) (n=5) and rapid recovery (“RAP”) (n=4)). Differential expression results relative to healthy subjects were compared for each pair of septic time points (left panel: day 4 vs CCI, middle panel: day 4 vs RAP, right panel: RAP vs CCI). The x-axis is the absolute difference in the *p*-value per gene (|Δ *p*-value|) and the y-axis is the difference in log fold-change (Δ logFC). The colored points represent genes that were differentially expressed in a single group or for both groups (*p*-value < 0.01). **(B)** Venn diagram of genes with overlapping significant differential expression (*p*-value < 0.01). **(C)** Enrichment results of significant genes representing the gene ontology biological processes. The y-axis is the negative log (base 10) of the *p*-value (- log_10_(pvalue)). **(D-F)** Similar to **(A-C)** for M-MDSCs. PMN: granulocytic, M: monocytic.

The greatest differences in the magnitude of M-MDSC gene expression from healthy subjects compared to septic patients occurred during acute sepsis (day 4) (**Fig. 2D**). In this cohort, 601 genes were differentially expressed in M-MDSCs, and only 31 of these were also significantly differentially expressed in late sepsis patients who either rapidly recovered or developed CCI at days 14-21 (**Fig. 2E)**. Gene ontology analysis among M-MDSCs revealed that patients experiencing rapid recovery had over-expression of kinase agents versus oxoacid metabolism when compared to sepsis patients with CCI (**Fig. 2F**).

In summary, transcriptomic analysis comparing healthy subjects, patients day 4 post-sepsis, and patients at days 14-21 post-sepsis (combining those who rapidly recovered with those who developed CCI) revealed significant heterogeneity in MDSC transcriptomics. In addition, lymphocyte suppressive activity, specifically suppression of T cell proliferation and T cell cytokine/chemokine production, varied between cohorts (*see below*). MDSCs from both time points after sepsis were dissimilar when comparing their gene expression profiles and significantly enriched biological processes from gene ontology.

### 3.2 Verifying the immunosuppressive capacity of MDSCs present in sepsis

Our laboratory has previously used cell sorting and subsequent T-cell suppression assays to demonstrate the immunosuppressive capacity of total MDSCs from septic patients (4, 16). Thus, we set out to verify that functionally active PMN- and M-MDSCs were indeed present in the isolated peripheral blood mononuclear cells (PBMCs) of septic individuals, as defined by Gabrilovich, et al (65). Surprisingly, we discovered unexpected cell types in our flow cytometry samples that were supported by single-cell analysis. Historically in the cancer literature, CD15 positivity is used to distinguish PMN-MDSCs from M-MDSCs (13). However, in representative septic patients, after identifying CD11b^+^CD33^+^ cells (**Fig. S1A**) and isolating HLA-DR^-/low^ cells to obtain the total MDSC population (**Fig. S1B**), CD15 was unable to clearly separate MDSC subpopulations (**Fig. S1C, bottom panel**). Alternatively, CD66b (*CEACAM8*; a granulocytic marker) was able to better delineate MDSC subpopulations (**Fig. S1C, top panel**) and, thus, was selected to distinguish PMN-MDSCs from PBMCs for functional analysis (**Fig. S1D**) (19, 20, 65-67). We undertook bulk CD66b^+^ cell isolation (STEMCELL Technologies, Vancouver); however, although CD14^-^CD15^+^ PMN-MDSCs enrichment was achieved, further analysis of the CD66b^+^-isolated cells demonstrated distinct CD66b^low^ and CD66b^high^ populations (**Fig. S2A**). Thus, we had enriched CD66b^low^CD14^+^CD15^-^ M-MDSCs in our gating strategy which was meant to only contain PMN-MDSCs (CD66b^high^) (**Fig. S2B**).

Functionally, the CD66b^+^ cells isolated from septic patient PBMCs suppressed either CD4^+^/CD8^+^ T-lymphocyte cytokine/chemokine production (**Fig. S3**) or lymphocyte proliferation of host CD8^+^ T-lymphocytes (**Fig. S4**), thereby meeting the criteria of MDSCs. CD66b^+^ MDSCs from septic but not healthy subjects altered T-cell cytokine production in response to antiCD3/CD28 treatment, including IFN-γ, IL-2, IL-4, IL-10, and IL-17 (**Fig. S3**). Cytokines which were analyzed which did not exhibit CD66b^+^ inhibition in acutely septic patients include IL-12, IL-23, and TGF-β. Only CD66b^+^ cells isolated from sepsis patients 14-21 days after infection were capable of significantly suppressing CD8^+^ T-lymphocyte proliferation in response to CD3/CD28 stimulation (**Fig. S4**). Although we did not see suppression of CD4^+^ T-lymphocyte proliferation by CD66b^+^ cells in response to CD3/CD28 stimulation, we did see a significant decrease in CD4^+^ T-lymphocyte proliferation stimulated in culture at days 14-21 compared to day 4 (**Fig. S5**). This indicates that CD4^+^ T lymphocytes are incapable of appropriate proliferation 2-3 weeks after sepsis (similar to what has been previously reported) (68), and that MDSCs at this time point may not be able to further suppress this aspect of CD4^+^ T lymphocyte function (22, 68).

### 3.3 Emergent view of MDSCs and transcriptomic analysis of a novel MDSC subpopulation

After our initial steps demonstrated that identification of MDSC subsets based on cell surface markers was potentially problematic in sepsis, we transitioned to cell classification via gene expression for the remainder of our analysis. All cells were clustered based on their transcriptomic profiles (visualized via UMAP (**Fig. 3A**) with relative percentages of each cell type depicted in **Fig. 3B**). The broad cell types were compared via expression of cell-surface marker genes (**Fig. 3C**) as well as percentage of spliced mRNA between patient groups (**Fig. 3D**). This was followed by careful manual annotation and inspection of canonical marker genes with identification of myeloid cells via differential expression of genes (**Fig. 4**). As explained by Hegde, et al. (13), there is substantial plasticity within MDSC subpopulations during sepsis which informs the relationship between MDSCs and terminally differentiated effector cells (**Fig. 5A**). Additional marker genes were used to obtain fine-level annotation of myeloid cell types (**Fig. 5B**). Importantly, four distinct populations of MDSCs were identified via this approach (**Fig. 5C**), three of which were consistent with classically defined E-, PMN-, and M-MDSCs (65). A novel fourth population was identified in 60% of the late sepsis patients who developed CCI, as well as both of the acutely septic patients who progressed to CCI (**Table 4**, **Fig. 6A**). This MDSC subpopulation exhibited gene expression patterns that were partially consistent with both M- and PMN-MDSCs. We thus labeled these cells "*hybrid*” (H)-MDSCs. Although one of the patients with CCI had a much greater number of H-MDSCs than other patients, it should be noted no H-MDSCs were observed in late sepsis patients who had rapidly recovered, or acutely septic patients who progressed to rapid recovery (**Table 4**).

**Figure 3.**
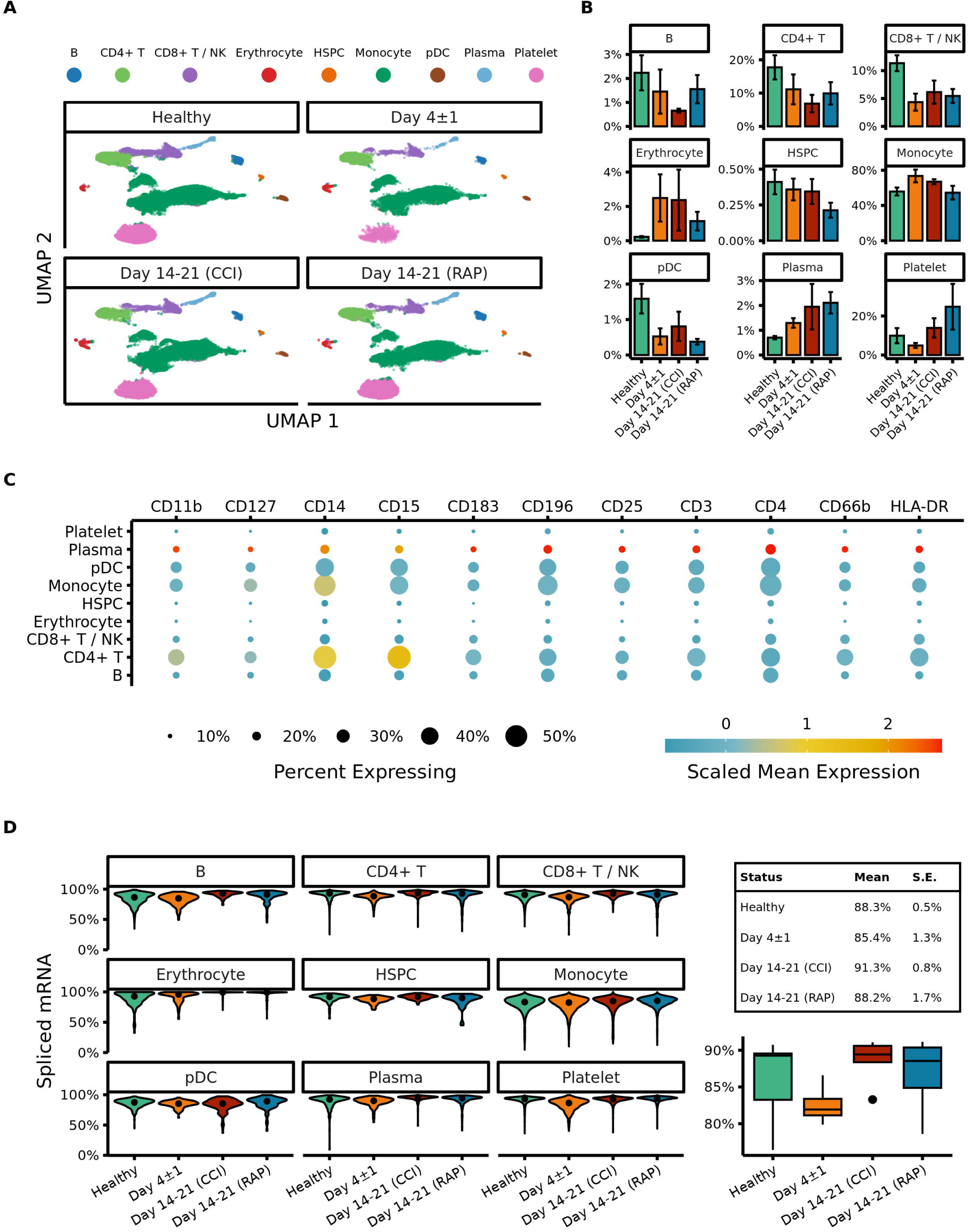
UMAP embeddings of peripheral blood mononuclear cells (PBMCs). **(A)** Cells are colored by the time point at which the samples were taken. Samples from acutely septic patients (“Day 4±1”) and late sepsis patients who either developed chronic critical illness (“CCI”) or experienced rapid recovery (“RAP”). **(B)** Cells are colored by septic cohort. Late sepsis patients at day 14-21 are separated into two groups denoted “CCI” (chronic critical illness) and “RAP” (rapid recovery) based on their response to the sepsis. **(C)** Expression of surface markers on subtypes of PBMCs. **(D)** The left panel denotes percentages of spliced mRNA in different cell types separated by patient cohort. The right panel denotes overall unspliced mRNA across cell types by patient cohort. B: B cells, NK: natural killer cells, HSPC: hematopoietic stem and progenitor cells, pDC: plasmacytoid dendritic cells.

**Figure 4.**
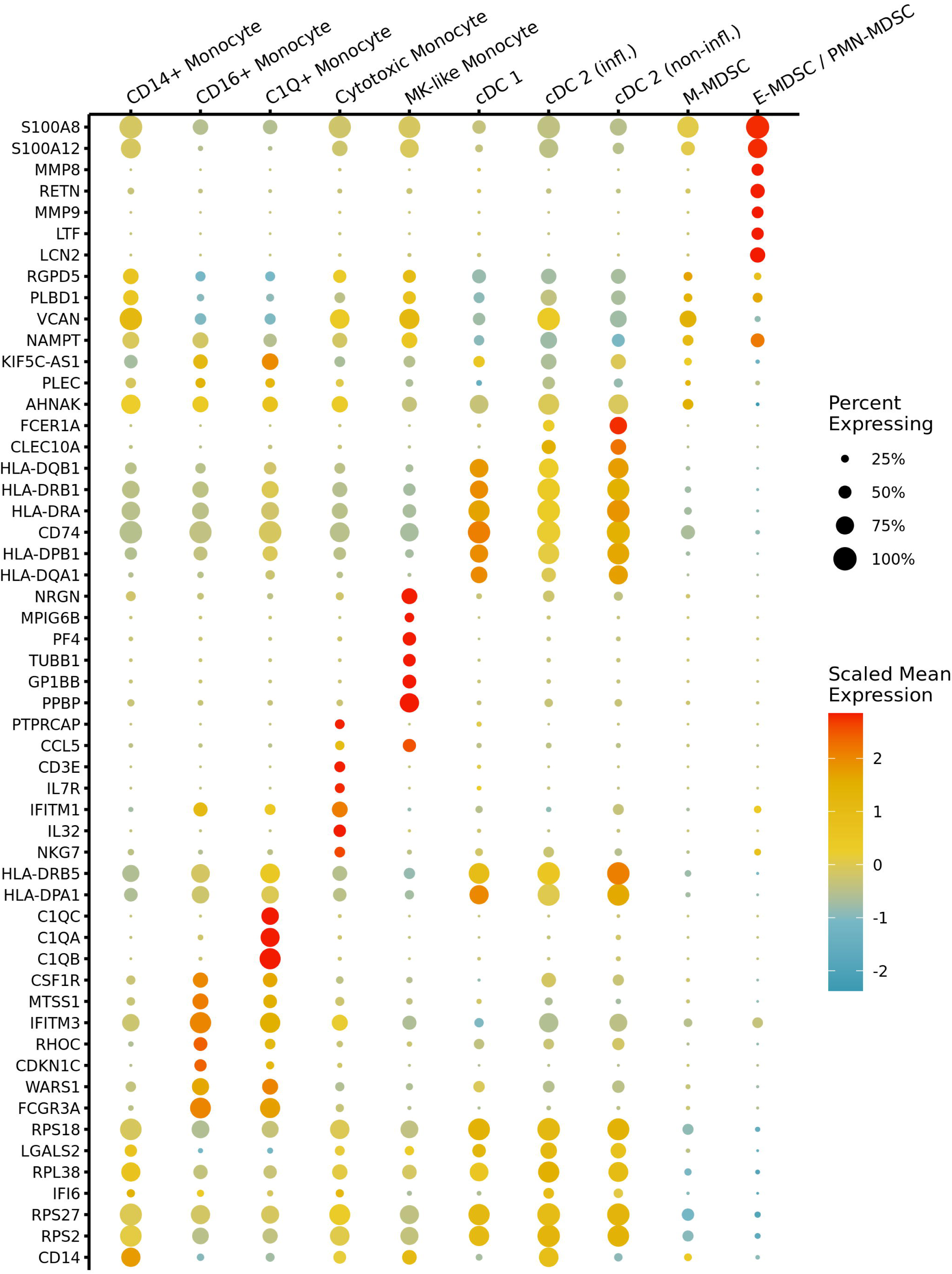
Marker gene expression across myeloid cell types in septic patients. A dot plot shows scaled mean expression of the top seven most significant differentially expressed genes (DEGs) in each myeloid cell type prior to fine-level annotation for MDSC subpopulations. Point radius indicates the percentage of cells with nonzero expression, and color denotes relatively higher or lower mean expression across cell types. Testing was performed with the Wilcox test, and genes were ranked by adj. *p*-value after Bonferroni correction. CD14^+^: classical monocyte, CD16^+^: non-classical monocyte, MK: megakaryocyte, cDC: conventional dendritic cells, infl.: inflammatory, M: monocytic, E: early, PMN: granulocytic.

**Figure 5.**
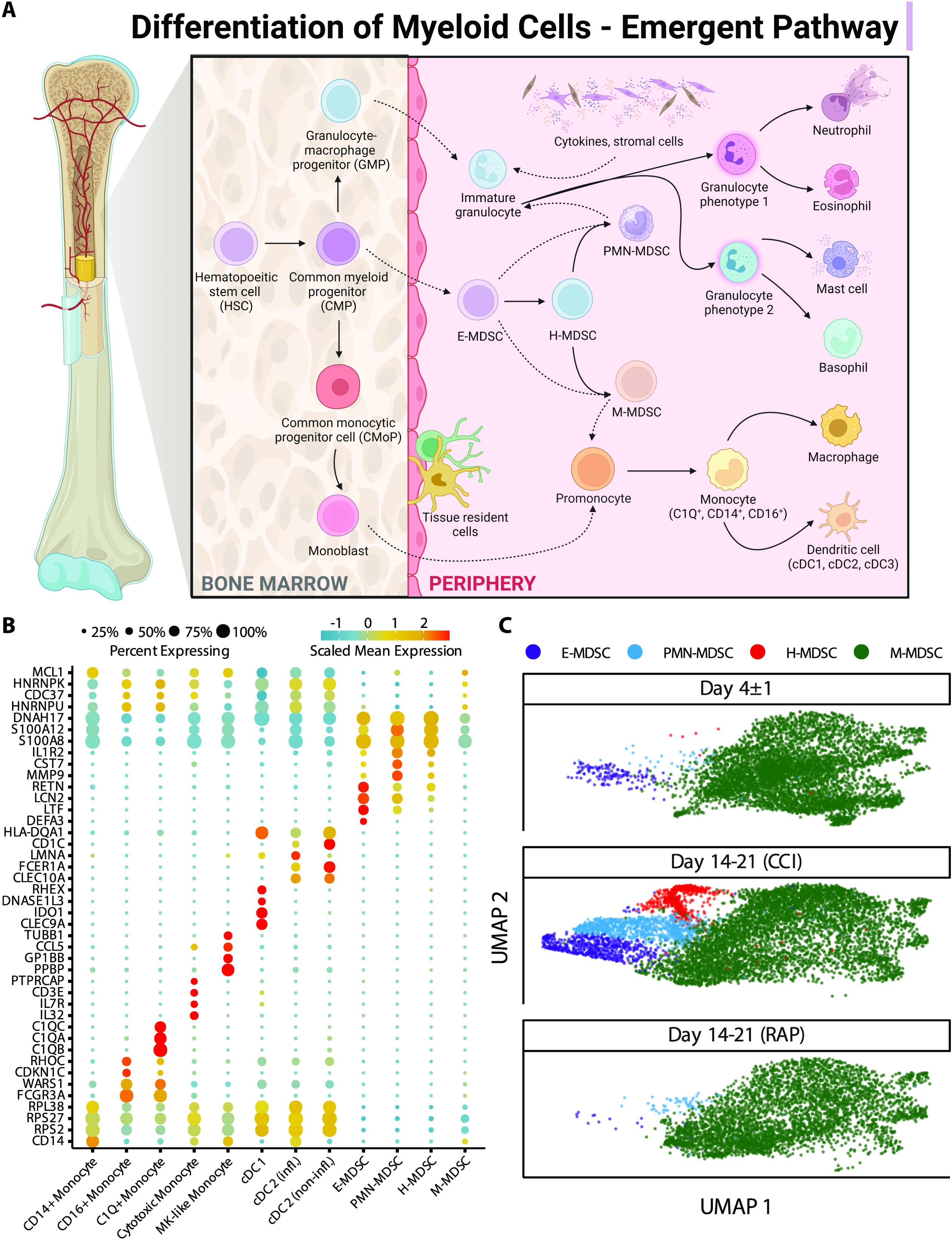
“Emergent” view and annotation of myeloid cell subpopulations in septic patients. **(A)** Illustration representing the “emergent” definition of MDSCs, incorporating the plasticity and heterogeneity of the myeloid compartment (modified from Hegde, et al (13)). **(B)** Fine cell type annotations within cells from septic patients that were broadly annotated as monocytes. The x-axis includes the different myeloid cell subtypes. The y-axis includes genes which were most highly expressed by each cell subtype. The scaled mean expression is denoted by the color of the dots, and the percentages of cells expressing the genes are represented by the size of the dots. **(C)** UMAP plots of cells of the four distinct subpopulations of MDSCs stratified by acutely septic patients (“Day 4±1”) (n=4), late sepsis patients who developed chronic critical illness (“CCI”) (n=5) or experienced rapid recovery (“RAP”) (n=4). This includes cells consistent with early (E-) MDSCs, granulocytic (PMN-) MDSCs, monocytic (M-) MDSCs, and a population of cells with characteristics of both M- and PMN-MDSCs, labeled hybrid (H-) MDSCs. MK: megakaryocyte, cDC: conventional dendritic cell, infl.: inflammatory.

**Figure 6.**
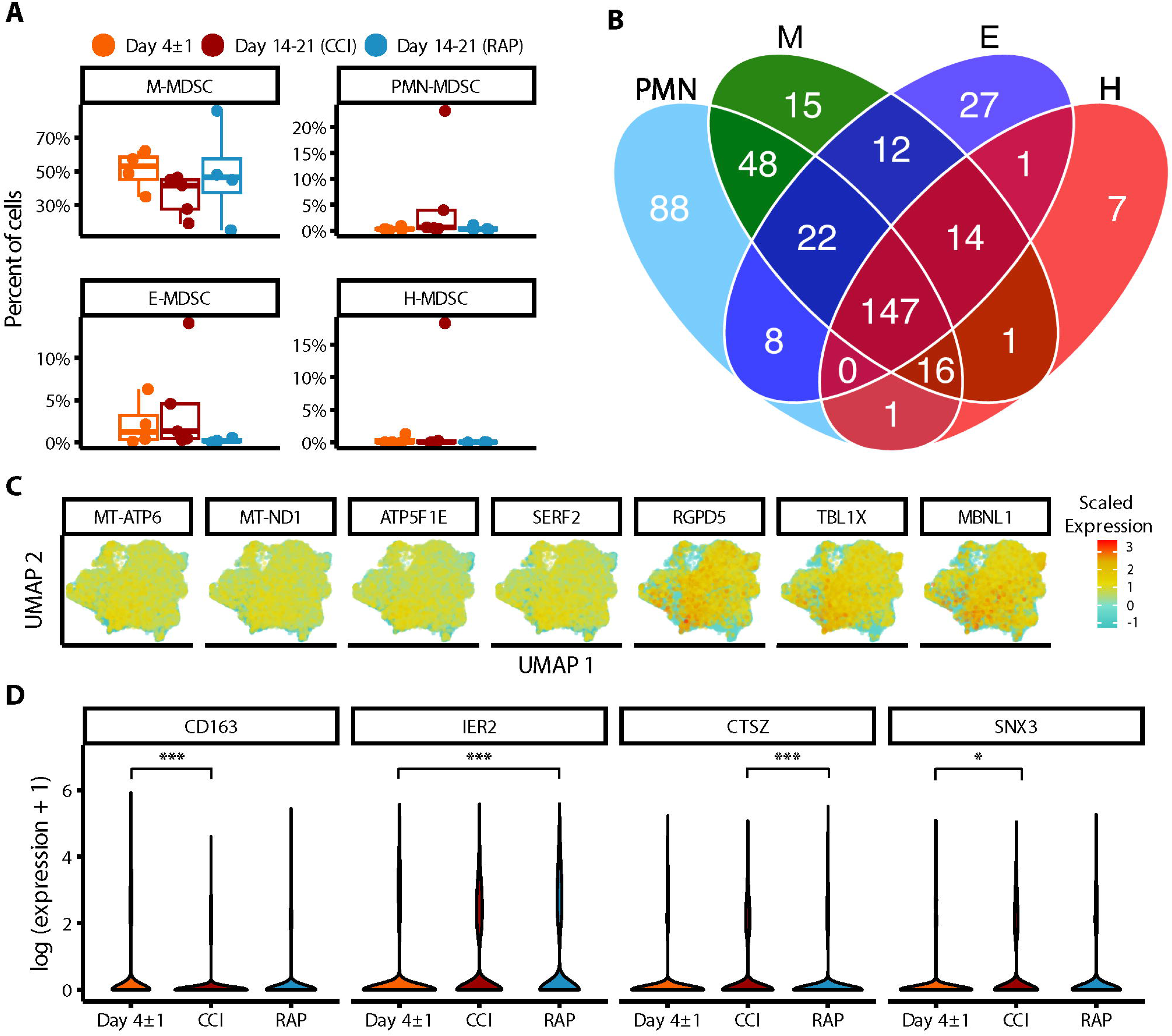
Characterizing data-driven subpopulations of MDSCs. **(A)** Relative frequencies of MDSCs by subpopulation. Percent of cells defined by transcriptomic analysis and gene expression, rather than cell surface markers. Grouped by acutely septic patients (“Day 4±1”) (n=4) and late sepsis patients who developed chronic critical illness (“CCI”) (n=5) or experienced rapid recovery (“RAP”) (n=4). **(B)** Diagram of significant marker genes for each MDSC subpopulation were determined in the pooled septic patients. **(C)** UMAP plots of all MDSCs are shown for the seven genes that were unique markers of gene expression in the H-MDSC subpopulation compared to all other MDSCs. Scaled expression represented by heat map of each gene. **(D)** Differential expression testing between septic groups in M-MDSCs revealed four genes that were significant. Y-axis is log (expression +1). Asterisks represent *p*-value cutoffs of 0.05 and 0.001, respectively, obtained from the mixed model analysis. M: monocytic, PMN: granulocytic, E: early, H: hybrid.

**Table 4.** H-MDSC cell counts by patient and associated outcome after sepsis. H-MDSC cell counts as determined by manual annotation. If blood samples were taken from acutely septic patients at day 4, then their eventual sepsis classification has been recorded. H: hybrid, CCI: chronic critical illness.

| Patient # | Patient Classification | Eventual classification if acute sepsis | H-MDSC Cell Counts |
| --- | --- | --- | --- |
| 1 | Acute Sepsis | Early death | 0 |
| 2 | Acute Sepsis | Rapid recovery | 0 |
| 3 | Acute Sepsis | CCI | 1 |
| 4 | Acute Sepsis | CCI | 8 |
| 5 | Rapid recovery |  | 0 |
| 6 | Rapid recovery |  | 0 |
| 7 | Rapid recovery |  | 0 |
| 8 | Rapid recovery |  | 0 |
| 9 | CCI |  | 552 |
| 10 | CCI |  | 6 |
| 11 | CCI |  | 1 |
| 12 | CCI |  | 0 |
| 13 | CCI |  | 0 |

The majority of MDSC-specific genes in septic patients were shared by at least two of the four subpopulations (66%, n=270 genes), with 36% (n=147 genes) significantly expressed by all four (**Fig. 6B, Supplemental File 2**). Although the MDSC subpopulations were fairly similar in terms of overlapping genetic expression, we identified seven genes uniquely expressed by H-MDSCs: *RGDP5, TBL1X, MBNL1, SERF2, ATP5F1E, MT-ND1,* and *MT-ATP6* (**Fig. 6C**). Gene expression was downregulated in RANBP2-like and GRIP domain-containing protein 5 (*RGDP5*), transducin (beta)-like 1X-linked (*TBL1X*), and muscleblind-like splicing regulator 1 (*MBNL1*) (69, 70). *TBL1X* regulates transcriptomic pathways and is upregulated in malignancy (69). *MBNL1* regulates alternative splicing and can be up- or downregulated depending on the type of cancer (71). Genes with upregulated expression included small EDRK-rich factor 2 (*SERF2*), ATP synthase F1 subunit epsilon (*ATP5F1E*), NADH-ubiquinone oxidoreductase chain 1 (*MT-ND1*), and mitochondrially encoded ATP synthase membrane subunit 6 (*MT-ATP6*). The latter three genes encode proteins involved in mitochondrial metabolism and function (72).

We next sought to identify differential genetic expression between our septic cohorts, specifically looking at differences between MDSCs in late sepsis patients who rapidly recovered and those who developed CCI. For differential expression across sepsis groups, only M-MDSCs were sufficiently present per group for fitting a linear mixed model with multiple subjects, as MDSCs are a relatively rare population overall. We identified four differentially expressed genes in M-MSDCs using this method: *CD163*, *IER2*, *CTSZ*, and *SNX3* (**Fig. 6D**). Expression of *CD163* (73), a gene responsible for controlling inflammation, was significantly lower in M-MDSCs in late sepsis patients with CCI versus acutely septic patients. *SNX3* has been identified as a potential septic biomarker (74), and was significantly upregulated in patients with CCI compared to acutely septic patients. *IER2* was significantly higher expressed in late sepsis patients who rapidly recovered compared to acutely septic patients. *IER2* is known to be upregulated in response to external stimuli including infection (75, 76). *CTSZ* expression was significantly higher in patients with CCI compared to patients who rapidly recovered after sepsis, and has been previously identified as a septic marker in mice (77).

The plasticity of the H-MDSC subpopulation is evident in the increased per-cell proportion of unspliced mRNA, indicating more active transcription. Only E-MDSCs had a higher proportion of unspliced mRNA in the myeloid compartment (**Fig. 7A**). To examine factors driving cellular activities, we identified genes with a high average proportion of unspliced mRNA within each cell subpopulation and performed enrichment analysis to identify relevant biological processes. Rather than focusing on individual ontologies, we used a network-based approach to cluster similar significantly enriched biological functions for each MDSC subpopulation (**Fig. 7B-E**) (52). Not surprisingly, actively transcribed genes in all MDSC subpopulations were enriched for activities pertaining to ‘*immune activation*.’ While PMN- and M-MDSCs had more biologically distinct functions, H-MDSCs shared enrichment with both cell types, specifically pertaining to pathophysiological septic-related processes including ‘*organonitrogen*’ and phosphorus-related processes.

**Figure 7.**
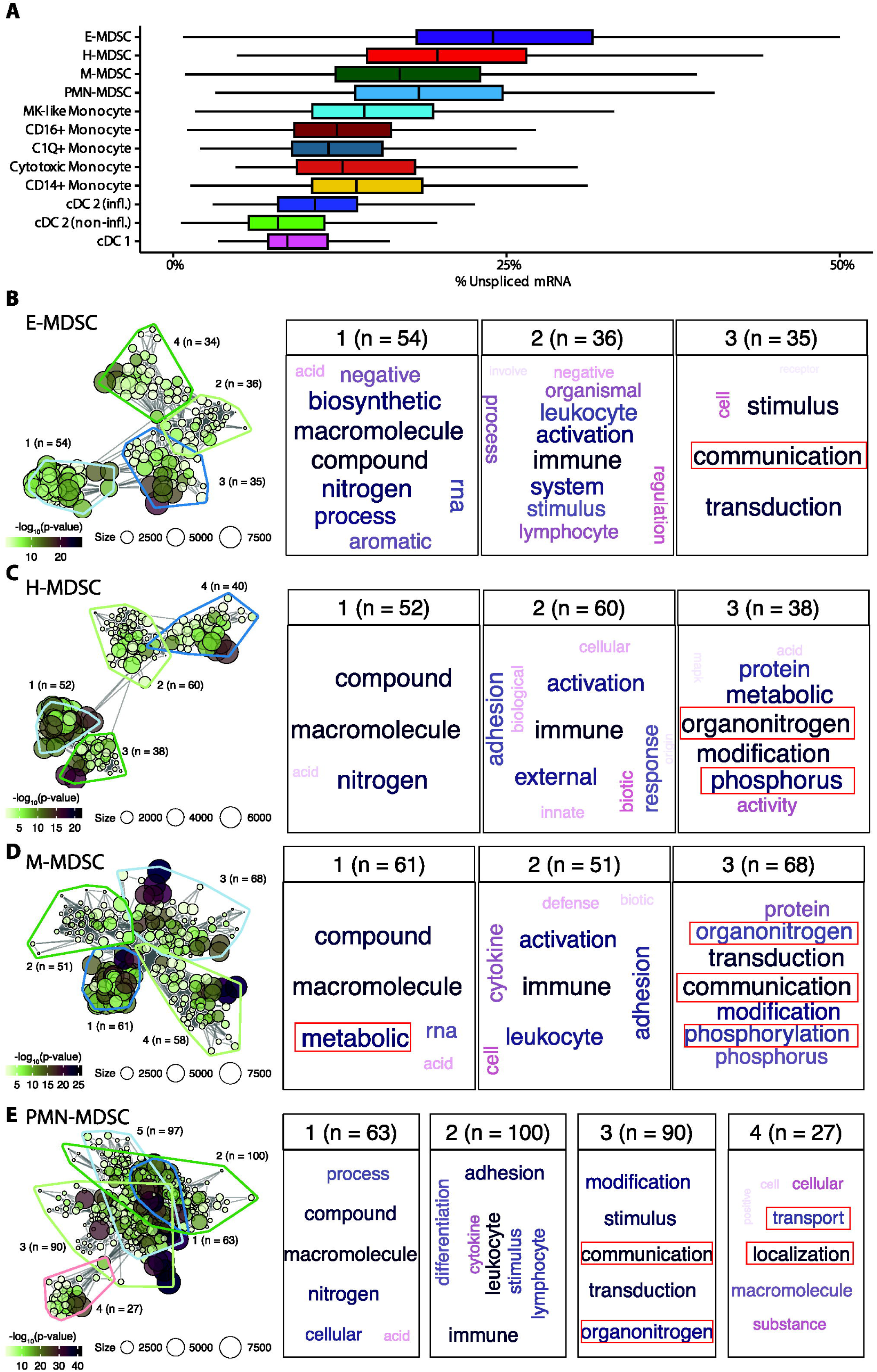
Larger proportions of unspliced mRNA in E- and H- MDSCs. **(A)** Distribution of unspliced mRNA percent across myeloid cell types. **(B-E)** Gene-set enrichment analysis of genes having high proportions of unspliced mRNA within each MDSC subpopulation. The left panel shows the gene-set network and clustering of significantly enriched biological processes. The right panels show word clouds for each biologically similar cluster (a general cluster of high-level biological processes was present for each cell-type and omitted). E: early, H: hybrid, M: monocytic, PMN: granulocytic, CD16^+^: non-classical monocyte, CD14^+^: classical monocyte, MK: megakaryocyte, cDC: conventional dendritic cell, infl.: inflammatory.

### 3.4 Determination of differentiation pathways and cell lineage in septic cohorts

Having described the MDSC subpopulations, we next set out to incorporate these findings into differentiation pathways of the myeloid compartment in septic patients. Quantifying transcriptional kinetics via RNA velocity estimation revealed complex, fluid relationships between MDSC phenotypes (**Fig. 8A**). As expected, M-MDSCs appeared to serve as the bridge between early immunosuppressive cell types and mature myeloid cells such as monocytes and conventional dendritic cells (cDCs) (**Fig. 8B**). As our analysis was based on PBMCs, it was not possible to compare the transition from MDSCs to mature granulocytes (PMNs). Estimating the graph connectivity between monocyte-lineage cell types allowed us to quantify the strength of each undirected relationship, and showed that MDSC subpopulations are both highly interconnected and much more internally similar to each other than they are to populations of terminally-differentiated myeloid cells (**Figs. 8B-C**).

**Figure 8.**
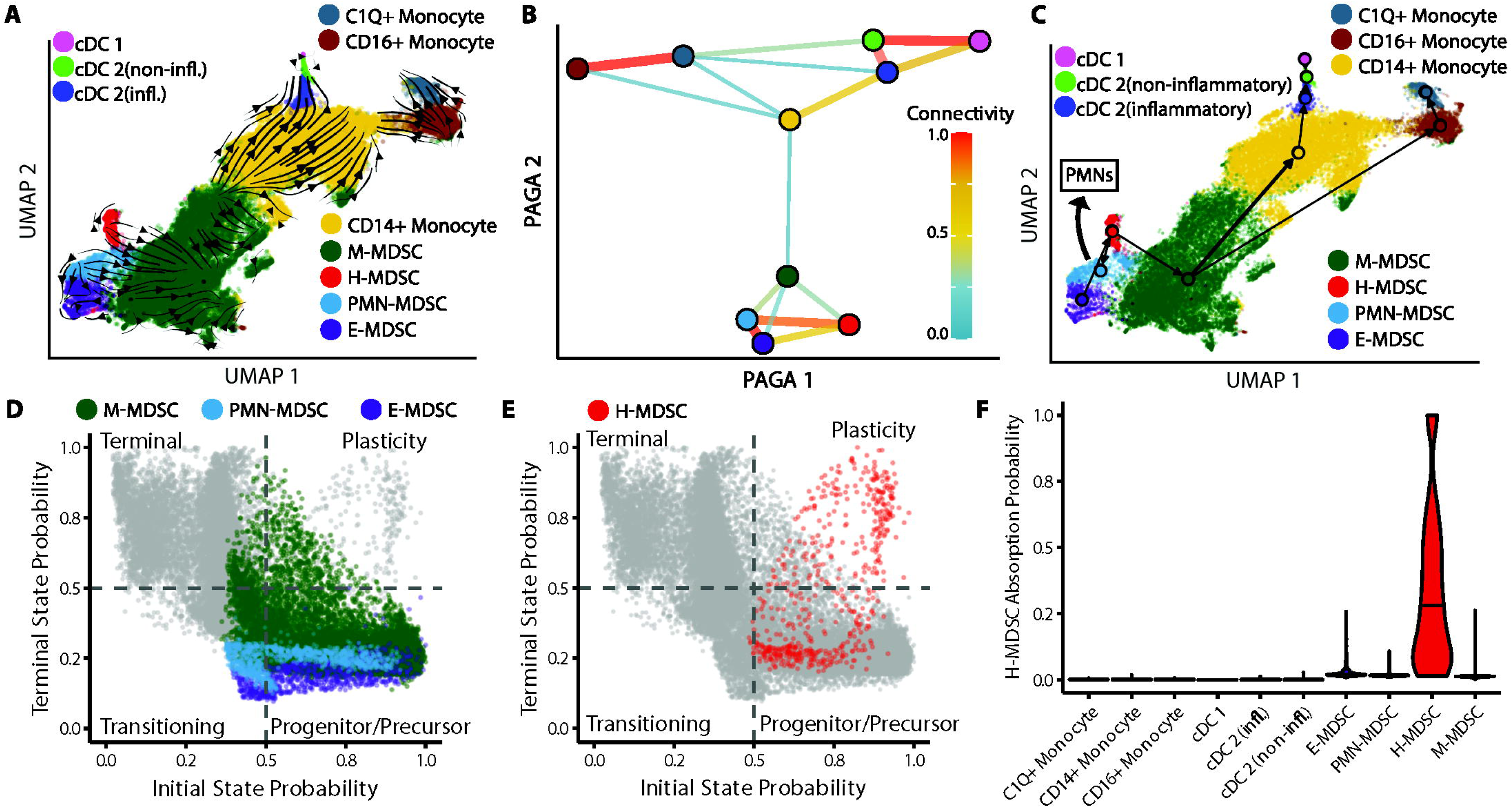
Topology of myeloid differentiation and plasticity in septic patients. **(A)** Myeloid cell smoothed RNA velocity estimates projected onto UMAP. Arrows represent differentiation potential. **(B)** Undirected partition-based graph abstraction (PAGA) of myeloid cell types. Line width/color between cell types denote relationship strength. Nodes colored by cell type. **(C)** Arrow directions represent differentiation potential. Arrow widths denote strength of connectivities between cell types. Arrow manually added indicating PMN-MDSC differentiation into granulocytes. **(D)** Cell state probabilities shown together for M-, PMN-, and E-MDSCs with all other cells in gray. **(E)** Similar to **(D)** with H-MDSCs in red. **(F)** H-MDSC cell fate absorption probabilities. cDC: conventional dendritic cell, infl.: inflammatory, CD16^+^: non-classical monocyte, CD14^+^: classical monocyte, M: monocytic, H: hybrid, PMN: granulocytic, E: early.

After analyzing velocity-inferred cell state transitions performed with **CellRank**, all E-, PMN-, and the vast majority of M-MDSCs cell states were classified as progenitor-like or transitioning-like (**Fig. 8D**). Only H-MDSCs contained a significant proportion of cells in a plasticity-like state with high probabilities for both initial and terminal cell states (in which cells remain H-MDSCs) (**Fig. 8E**) (57). Supporting this, significant variation was observed in the likelihood of an H-MDSC staying an H-MDSC when estimated by absorption probabilities from **CellRank**, with a mean (SD) probability of 0.33 (0.29) (**Fig. 8F**). No other cell types were likely to end up as H-MDSCs. To better characterize the biology underlying commitment to the H-MDSC cell fate, lineage driver genes (genes significantly correlated with the probability of becoming an H-MDSC) were identified by computing Spearman correlations of expression with absorption probabilities. Highly correlated genes were diverse in function and included inflammation-associated genes such as *S100A8*, *-9*, and *-12*, along with immunoregulatory genes *ALOX5A*, *RETN*, and *IL1R2*.

Next, we investigated differences in cell states across sepsis groups for each MDSC subpopulation. As H-MDSCs were not observed in sepsis patients who experienced rapid recovery, they were not included for this analysis. M-MDSCs were highly consistent between septic patients at day 4 and days 14-21 in terms of their cell states and kinetics (**Fig. 9A**). Interestingly, PMN-MDSCs displayed the most heterogeneity, specifically in late sepsis patients with CCI compared to both day 4 septic patients and late sepsis patients who rapidly recovered. PMN-MDSCs in late sepsis patients who developed CCI had significantly slower differentiation speed, higher cell state stability, and lower initial state probabilities (**Fig. 9B**). This is consistent with PMN-MDSCs persisting in CCI compared to patients who rapidly recover after sepsis. E-MDSCs in late sepsis patients with CCI also showed significantly lower differentiation progression than acutely septic patients or late sepsis patients who rapidly recovered, along with a higher degree of cell commitment along the differentiation trajectory compared to acutely septic patients (**Fig. 9C**).

**Figure 9.**
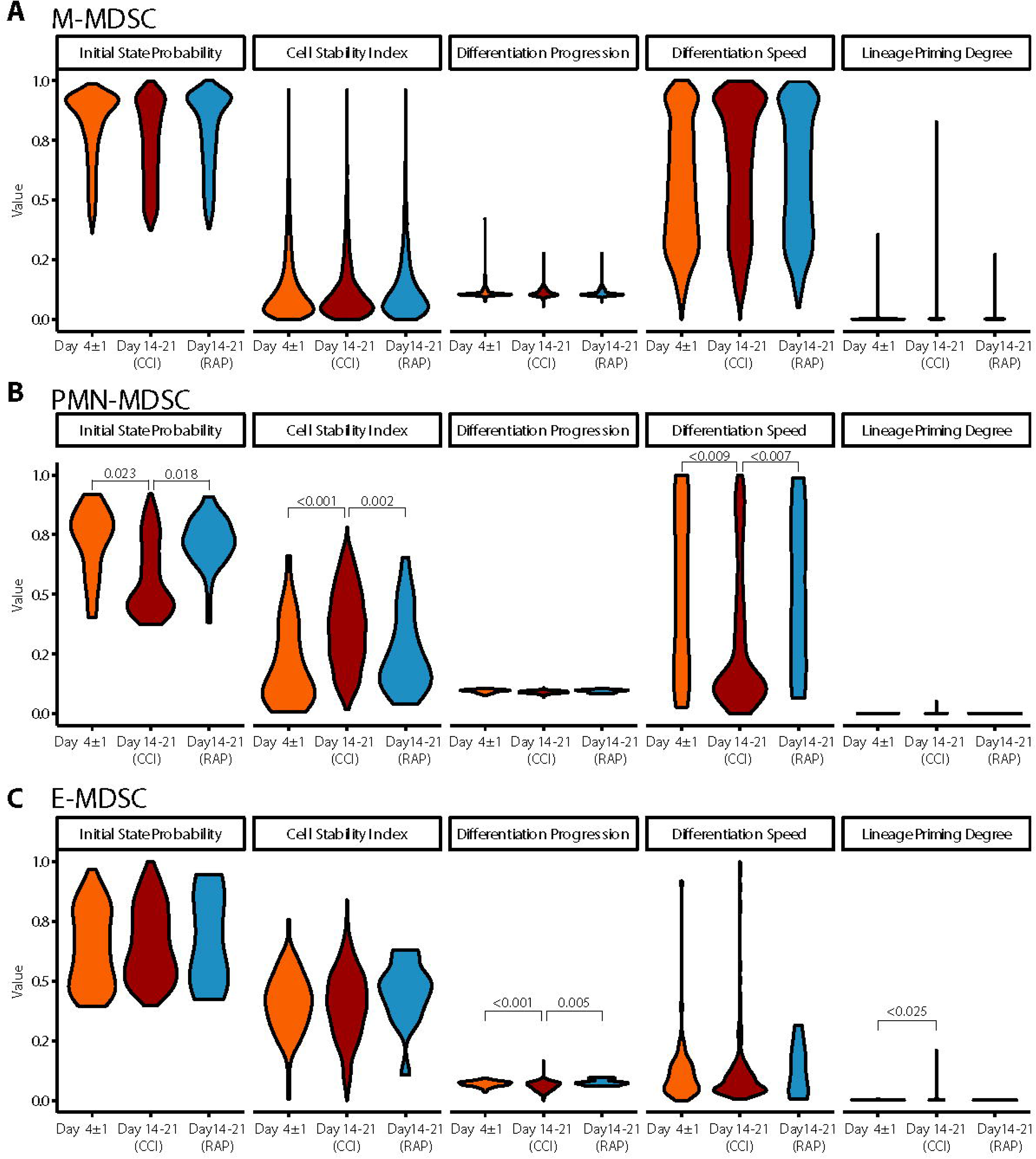
Differences in PMN-, E-, and M-MDSCs across septic time-points. **(A)** Cell dynamic parameters estimated from CellRank were compared across cells from septic patients at Day 4±1 (acute sepsis) (n=4), patients at day 14-21 who rapidly recovered (“RAP”) (n=4), and patients at day 14-21 who developed chronic critical illness (“CCI”) (n=5) in M-MDSCs. **(B-C)** Similar to **(A)** for PMN-MDSCs and E-MDSCs, respectively. Significant p-values (< 0.05) were obtained from fitting a linear mixed model. E: early, PMN: granulocytic, H: hybrid, M: monocytic.

As previously stated, CD66b^+^-isolated PBMCs met the criteria of MDSCs in their ability to suppress either T-lymphocyte cytokine/chemokine production or T-lymphocyte proliferation *ex vivo* (**Figs. 5 & 6**) (4, 16), although CD66b^+^-isolated PBMCs were not identical in their suppressive activity from acutely septic patients or late time periods after severe infection. Interestingly, whether using cell-surface markers or transcriptomic analysis of the current dataset, differential expression of several key MDSC genes published in the cancer literature did not reach significance and/or were modestly expressed in septic individuals (**Fig. 10**). For example, although there was upregulation of genes in the *S100A* and *MMP* superfamilies, differential expression of *ARG1, IL-10, NOS2,* and *TGFB1* did not reach significance (although transcripts from all genes were detected).

**Figure 10.**
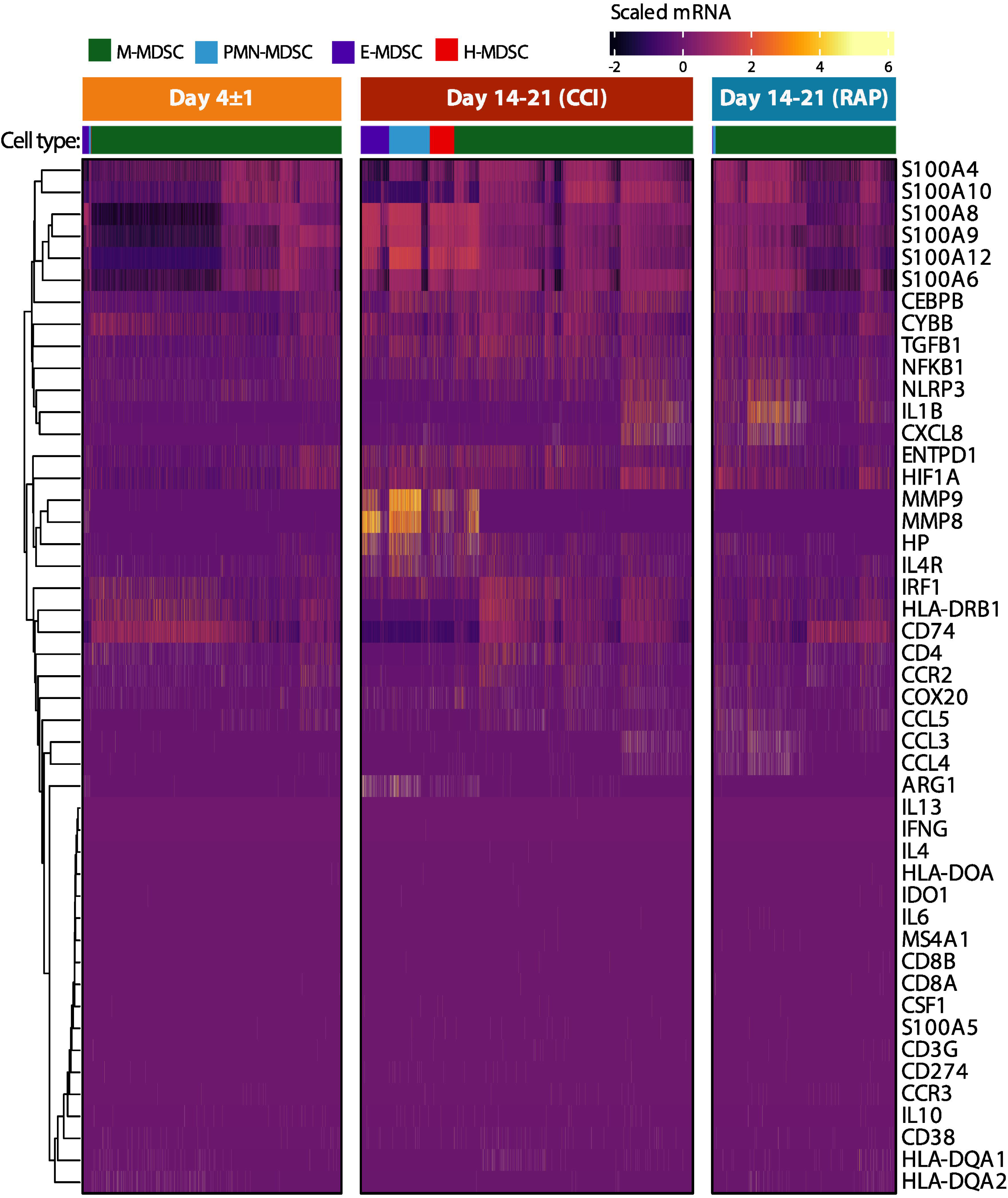
Canonical MDSC genes in immunosuppressive cell subpopulations in septic patients. Heatmap of scaled expression of canonical genes identified in the current MDSC literature. Cells in the four identified MDSC subpopulations are denoted in the colored key. Genes were arranged using hierarchical clustering with complete linkage. Patient groups include acutely septic patients (“Day 4±1”) (n=4) and late sepsis patients who developed chronic critical illness (“CCI”) (n=5) or experienced rapid recovery (“RAP”) (n=4). M: monocytic, PMN: granulocytic, E: early, H: hybrid.

## 4 Discussion

Since their delineation by Gabrilovich in 2007 (78), MDSCs have been reported in multiple inflammatory diseases in addition to cancer (79). Recently, Hedge et al. described significant heterogeneity among these immune suppressive cells in the myeloid compartment (13). They stated that historically we have had a ‘*monolithic view*’ or definition of MDSCs, and that a more complex ‘*emergent view’* is required to better understand these leukocytes (13). In this report, we have taken both conceptual approaches (monolithic and emergent) to analyze MDSCs in one of the first cohorts to compare patients with poor (CCI) versus good (rapid recovery) clinical outcomes after surgical sepsis. Importantly, all analyses revealed significant alterations in the evolution of MDSCs after sepsis (i.e. time points) as well as significant differences in the MDSC subpopulations taken from sepsis survivors who rapidly recovered or developed CCI. In classifying MDSCs via gene expression and transcriptomic analysis, we have also identified a novel MDSC subpopulation (H-MDSCs) present only in sepsis survivors with CCI and acutely septic patients who progressed to CCI. Finally, even though we have demonstrated in this work and previously (16) that these cells suppressed lymphocyte proliferation to antigenic stimulation (similar to oncologic processes), the MDSCs identified after sepsis do not significantly express many of the well-described genes key to MDSC immunosuppression in other pathologies, most commonly cancer (80).

The study of MDSCs has expanded dramatically over the past decade. However, the overwhelming majority of these studies performed using blood samples are from cancer patients; only five studies focus on systemic infection and sepsis (8, 14, 81-83). Although MDSCs are commonly detected in different inflammatory pathologies, there is a gap in research regarding this cell type in the infected or post-infected host. Data are increasingly illustrating the impact of a dysregulated myeloid compartment in patients with poor long-term outcomes, including COVID-19 (84). MDSCs have been identified in these patients, especially those with more severe disease or poor outcomes (85, 86), and are being considered as a target for immunotherapy (87).

MDSCs are challenging to define and characterize. As such, cell surface markers and genes historically used to identify MDSC subpopulations were amassed from multiple different resources, predominantly from the cancer literature. Surface markers differ between humans and other species, so only human studies could be considered (88, 89). Based on previous work, we began by isolating CD66b^+^ PBMCs as a means to obtain PMN-MDSCs for functional analysis in septic patients and healthy subjects (10). Interestingly, although we found that the purity of the isolation of CD66b^+^ leukocytes (**Fig. S1**) was very good, and even though CD66b is considered a marker for granulocytes (90), we identified that the CD66b^+^ population consisted of a mixture of PMN- and M-MDSCs (**Fig. S2**). Although there can be populations of MDSCs that have different levels of both CD14 or CD15 cell surface expression (91), these positively isolated CD66b^+^ PBMCs were a combination of CD14^+^CD15^-^CD66b^low^ (M-MDSC) and CD15^+^CD66b^high^ (PMN-MDSC) cells. Our CITE-seq data confirmed that *CEACAM8* expression was present in multiple myeloid cell populations. This variable MDSC cell surface expression of CD66b in septic patients appears similar to a cell type described in 1998 to define asynchronous myelopoiesis in malignant myeloid disorders (92). This highlights some of the difficulty regarding the use of cell surface phenotypes to classify MDSC subtypes after sepsis.

Of note, MDSCs are continuing to be described in certain patient populations, including sepsis, through cell surface markers only (93, 94). Although these data may be valid, our analysis would indicate that the traditional “*monolithic*” definition of MDSCs may not adequately define these plastic, transitory cell populations in critically ill patients with sepsis. Our results do not refute any currently accepted definitions of human MDSCs (including by cell marker phenotype) (10), but rather illustrate the complexity of the myelodysplasia that occurs after human sepsis, and the shortcomings of cell surface markers alone to identify myeloid cell types after severe infection. In addition, other immunosuppressive cells exist in the PBMCs of whole blood from septic human patients, specifically low-density PMNs and exhausted monocytes (95, 96). This work, and our current results, indicate an immediate compelling need for more refined and nuanced descriptions and definitions of the myeloid compartment after sepsis.

Veglia et al. previously described transcriptomic differences between MDSCs and terminally differentiated monocytes and neutrophils (66). Additional guidelines for characterization and nomenclature of MDSCs based on cell surface phenotypes have been proposed, although the same central resource does not appear to exist for single-cell transcriptomic signatures of different MDSC subpopulations (65, 97). However, specific genes have been described in the literature. In cancer, *STAT3* is important for the T-cell suppression exerted by MDSCs (98). *STAT1*, *-5*, and *-6* are also important in the regulation of arginase activity, although this may be more pertinent for cancer than sepsis based on the subdued level of *ARG1* expression in MDSCs identified from our septic patients (**Fig. 10**) (98). It should also be noted that different subpopulations than the canonical PMN- and M-MDSCs have been previously described, including Eo-MDSCs (with eosinophilic characteristics) and fibrocystic MDSCs (99, 100).

A population of H-MDSCs was found when using the “*emergent*” classification system of MDSCs via genetic expression in order to classify cell types (**Fig. 5C**). All four MDSC subpopulations appeared strongly interrelated and our data indicated that these cells are likely plastic in their myeloid state after sepsis (**Fig. 6B**) (13). As to why we classified these cells as unique from previously defined MDSC subpopulations, H-MDSCs express many similar genes as PMN-MDSCs, although the average expression of these genes tends to be lower, such as with *IL1R2, CST7* and *MMP8/9*. H-MDSCs also share substantial overlap with M-MDSCs, with higher expression in sepsis of genes like *S100A8/9* and *DNAH17* (**Fig. 10**). H-MDSCs may be an intermediary between MDSC subpopulations, and their presence in CCI further reveals the plasticity of myeloid differentiation in sepsis (**Fig. 8**). Although MDSC subpopulations share a similar phenotype after sepsis, their function and transcriptomic patterns are distinct. Thus, after sepsis, *‘a MDSC is not a MDSC,’* and there is a unique expression of myelodysplasia after severe infection depending on both host and outcome. These data support the concept that targeted therapeutic strategies will be required within these sepsis phenotypes given the heterogenic response of the myeloid compartment to sepsis.

This study was limited by the number of patients in each study arm; however, our sample size estimates were similar to past publications in the field (14, 101). This is also a single-institution study in which treatment of sepsis is standardized but may differ compared to other institutions. Additionally, we did not stratify septic patients by septic source. Future directions include stratification of our patient cohorts by infection source and demographic information such as age, sex, and ethnicity/race to determine confounding factors which may have affected our analysis by different clinical outcomes after sepsis.

In summary, we have determined that the post-septic myeloid compartment is complex and includes a unique MDSC subpopulation that has not been previously described. Importantly, the heterogeneous response of the blood myeloid compartment to sepsis varies based on time and clinical outcome (CCI vs rapid recovery) and demonstrates that cell surface markers may not be a reliable indicator of circulating myeloid cell types after sepsis. Sepsis, like many other pathologies, requires a precision/personalized medical approach in order to improve host outcomes (22). Our work reveals specific cell types and pathways that could be modified in patients at risk of poor outcomes after sepsis (CCI) to convert them to a phenotype of rapid recovery.

## 9 Author Contributions

Conceptualization: MK, CM, LLM, PAE

Methodology: LLM, PAE

Investigation: DBD, JCR, MW, MLD, RU, DCN, MLG, SL, LM, TL, AMM, RM, MK, CM, MAB, TMB, LLM, RB, PAE

Visualization: LLM, PAE

Funding acquisition: MK, CM, LLM, PAE

Project administration: PAE

Supervision: PAE

Writing – original draft: ELB, JL, VP, LLM, RB, PAE

Writing – review & editing: ELB, JL, DBD, JCR, MW, VP, GG, JM, MLD, RU, DCN, MLG, SL, LM, TL, AMM, RM, MK, CM, MAB, TMB, LLM, RB, PAE.

## 10 Funding

This work was supported, in part, by the following National Institutes of Health grants:

National Institutes of Health grant RM1 GM139690 (LLM, PAE, MK, CM)

National Institutes of Health grant R35 GM140806 (PAE)

National Institute of General Medical Sciences grant R35 GM146895 (RB)

National Institute of General Medical Sciences postgraduate training grant T32 GM-008721 (EB, DBD, VP, JM)

National Institute of General Medical Sciences postgraduate training grant T32 HL160491 (GG)

### 11 Acknowledgments

The authors would like to thank LaShaun Bryant, BS, Brandi Buscemi, AS - Physical Therapist Assistant, Ruth Davis, BSN, Jennifer Lanz, MSN, RN, Ashley McCray, ASN, and Ivanna Rocha, MPH for their critical role with patient recruitment, retention and data collection as well as collection of human samples.

## 12 Data availability

The datasets generated for this study can be found in the Gene Expression Omnibus (in-process). All analysis codes are available upon request.

