## Supplemental Figures for "The Post-Septic Peripheral Myeloid Compartment Reveals Unexpected Diversity in Myeloid-Derived Suppressor Cells"

Supplementary Material

### 1 Supplementary Figures

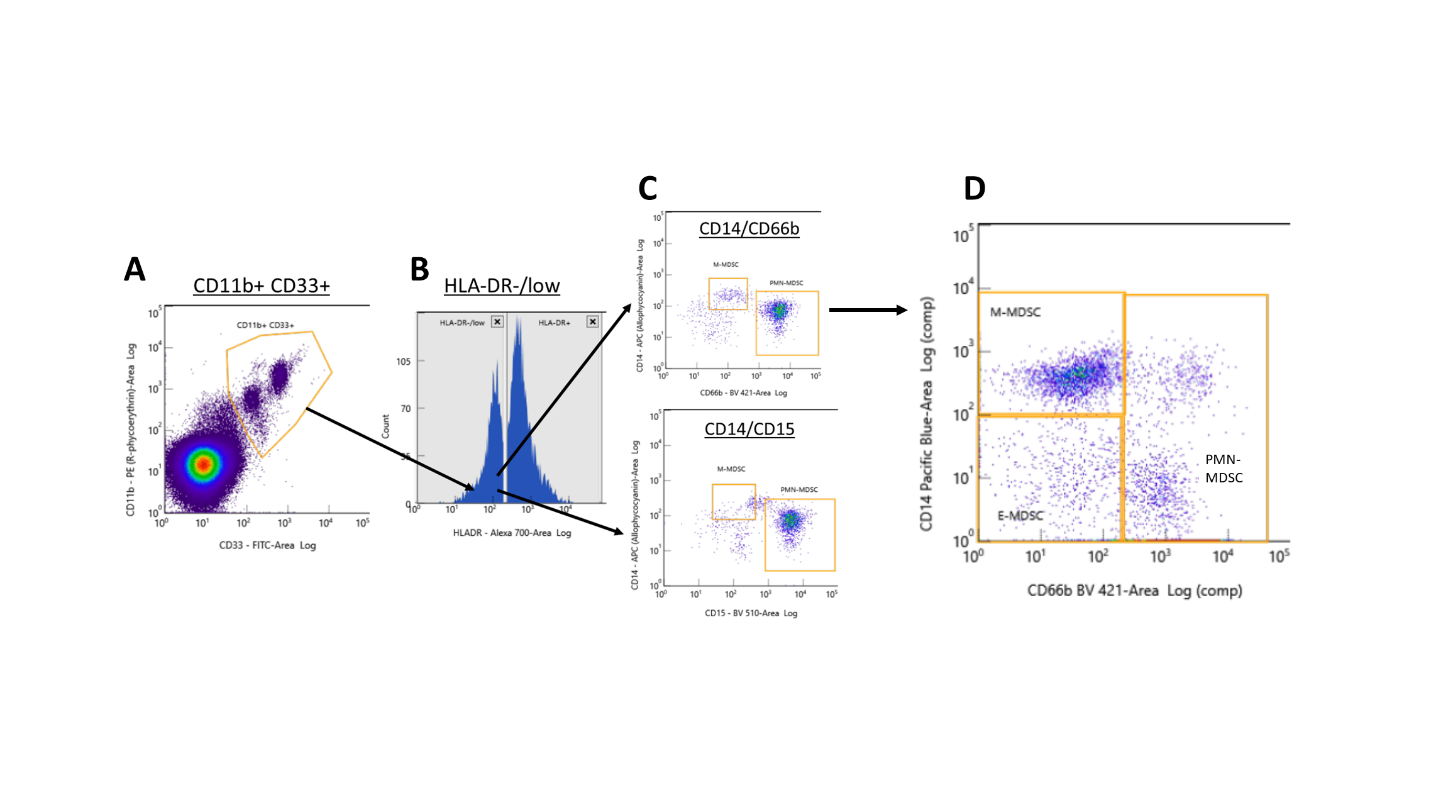

**Supplementary Figure 1. Identification of MDSC phenotypes via flow cytometry with the cell surface markers CD14, CD15, and CD66b.** **(A)** Cells were initially gated for doublet exclusion, then viable cells were determined using Sytox Live/Dead stain. Subsequently, the CD11b^+^ and CD33^+^ cells were gated. **(B)** HLA-DR^low^ cells were selected to capture the total MDSC population (CD11b^+^ CD33^+^ HLA-DR^low^). **(C)** Initially, CD14 and CD15 were used to isolate MDSC subpopulations (bottom panel); however, it was found that CD66b selection provided better discrimination (top panel). **(D)** From the total MDSC population, the E-MDSC (CD14^-^ CD15^-^ CD66b^-^), M-MDSC (CD14^+^ CD66b^low^), and PMN-MDSC (CD15^+^ CD66b^high^) subpopulations are determined.

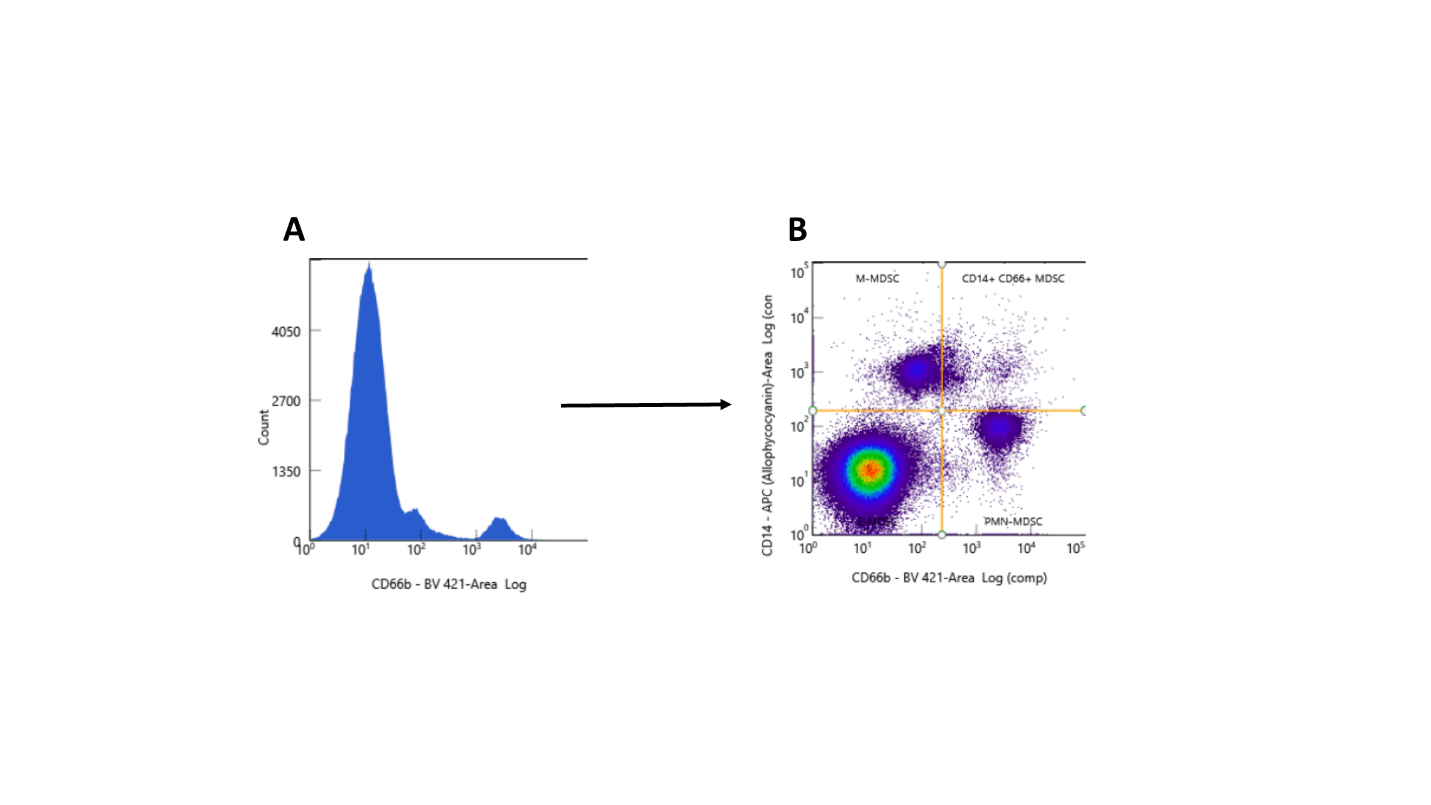

**Supplementary Figure 2. Isolation of CD66b^+^ cells from the PBMC layer in a septic patient. (A)** CD66b^+^ cells were isolated from the peripheral blood mononuclear cell (PBMC) layer via STEMCELL positive selection kit. Populations of CD66b^low^ and CD66b^high^ were visualized. **(B)** M-MDSCs are CD66b^low^ and PMN-MDSCs are CD66b^high^. Both were present in the CD66b^+^ isolation, which should just be PMN-MDSCs (CD66b^high^).

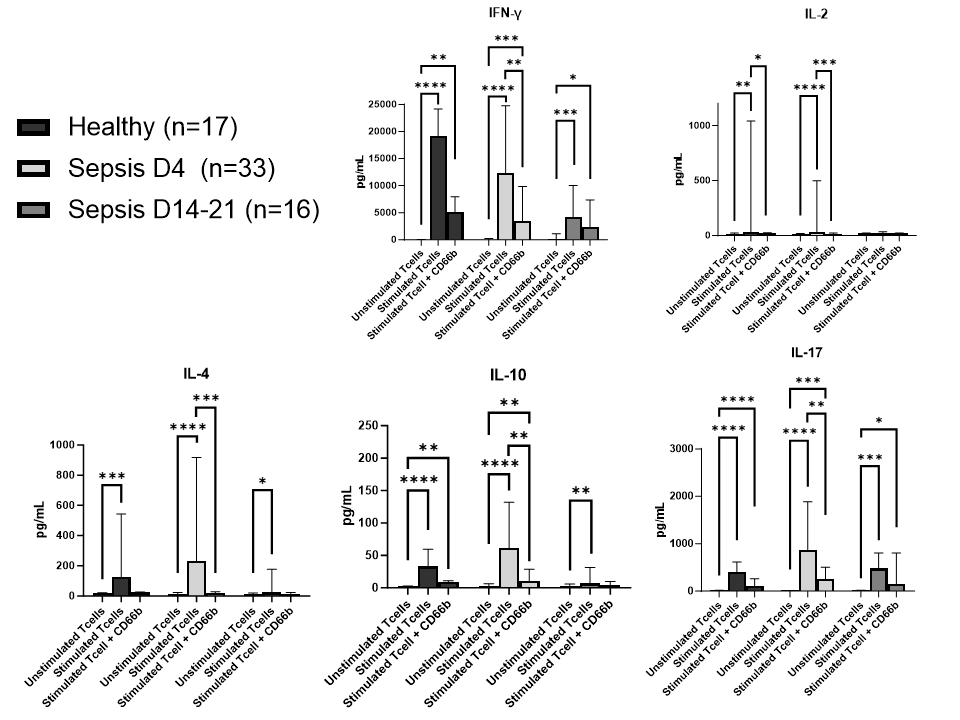

**Supplementary Figure 3.** **Proliferation index and cytokine production of CD4^+^ and CD8^+^ T cells in septic patients and healthy subjects.** Level of cytokine production by CD4^+^ and CD8^+^ lymphocytes in 10 healthy subjects (“Healthy”), 18 acutely septic patients (“Sepsis D4”), and 8 septic patients at day 14-21 (“Sepsis D14-21”). T lymphocytes plated and stimulated with soluble anti-CD3/CD28 antibodies or without (control). CD66b^+^ cells also isolated from the PBMC fraction via positive selection kit. Cells co-cultured with stimulated T cells in a 1:1 ratio. Supernatants after culture were obtained for cytokine analysis. The Kruskal-Wallis test was used for analysis of medians between groups. CD4: CD4^+^ lymphocytes, CD8: CD8^+^ lymphocytes. *: *p*<0.05, **: *p*<0.01, ***: *p*<0.001, ****: *p*<0.0001.

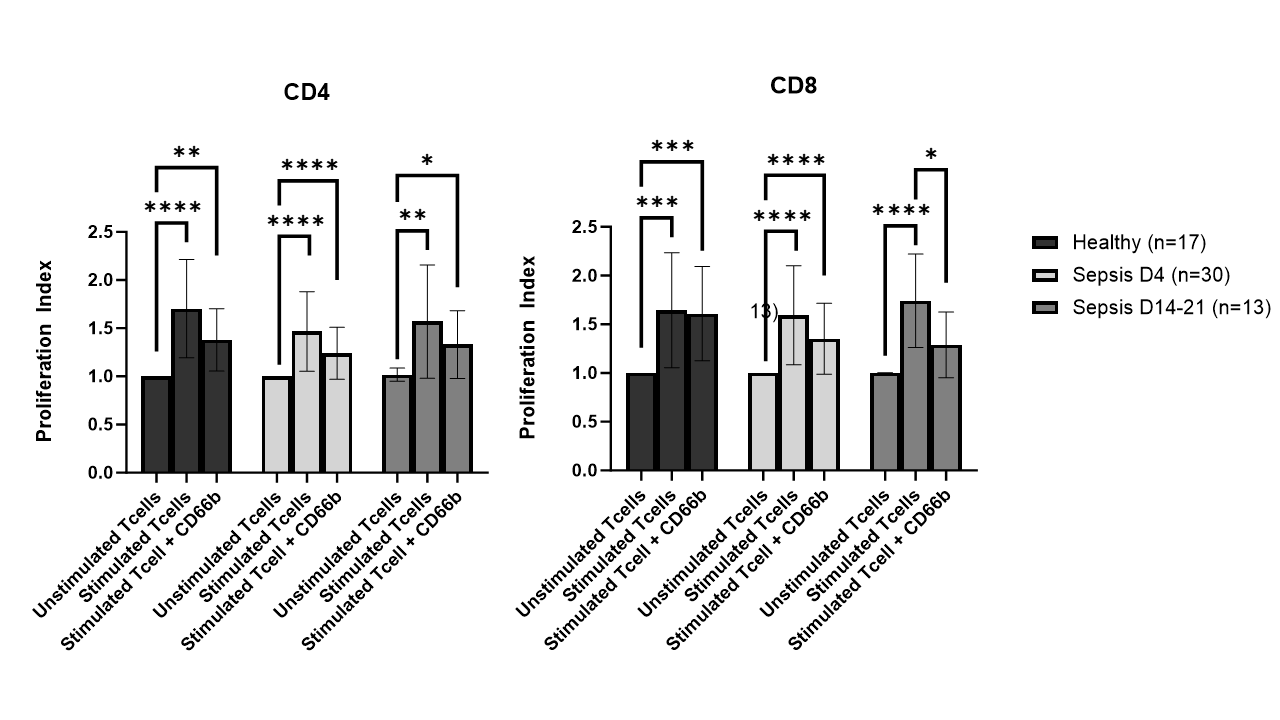

**Supplementary Figure 4.** **Proliferation index of CD4^+^ and CD8^+^ T cells in septic patients and healthy subjects.** T cells isolated from peripheral blood mononuclear cell (PBMC) suspension from 10 healthy subjects (“Healthy”), 18 septic patients at day 4 (“Sepsis D4”), and 7 septic patients at day 14-21 (“Sepsis D14-21”) using the same techniques as patients whose samples underwent processing and scRNA-seq. T lymphocytes plated and stimulated with soluble anti-CD3/CD28 antibodies or without (control). CD66b^+^ cells also isolated from the PBMC fraction via positive selection kit. Cells co-cultured with stimulated T cells in a 1:1 ratio. Proliferation indices calculated as the total number of cell divisions divided by the number of cells that went into division. The Kruskal-Wallis test was used for analysis of medians between groups. CD4: CD4^+^ lymphocytes, CD8: CD8^+^ lymphocytes. *: *p*<0.05, **: *p*<0.01, ***: *p*<0.001, ****: *p*<0.0001.

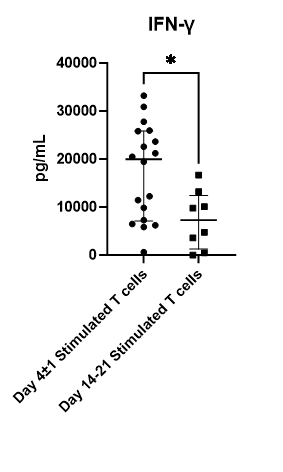

**Supplementary Figure 5.** **IFN-γ expression in stimulated T cells between acute and sub-acute sepsis patients.** CD4^+^ and CD8^+^ T cells were isolated from PBMC suspension. They were stimulated with soluble anti-CD3/CD28 antibodies and cultured. After 4 days, cells were harvested and supernatants analyzed for expression of IFN-γ. Groups were stimulated T cells from patients with acute sepsis at day 4 (n=18) and stimulated T cells from patient samples 14-21 days after sepsis onset (n=8). Difference assessed via unpaired non-parametric Mann-Whitney test. *: *p*<0.05.
